# Cell type-specific placental signatures in polyendocrine metabolic ovary syndrome and metformin

**DOI:** 10.64898/2026.05.21.26353338

**Authors:** Hong Jiang, Xuelei Wang, Eszter Vanky, Denise Parreira, Emilie Derisoud, Paulo R Jannig, Emelie Nordenhök, Allan Zhao, Congru Li, Solhild Stridsklev, Malin Holzmann, Xinmin Li, Charlotte Millde Luthander, Elisabet Stener-Victorin, Qiaolin Deng

## Abstract

Polyendocrine metabolic ovarian syndrome (PMOS) is associated with adverse pregnancy outcomes and increased cardiometabolic risk in offspring, yet the underlying placental mechanisms remain poorly understood. Metformin is widely prescribed in these pregnancies despite limited mechanistic justification. Here we apply multimodal molecular analyses of placentas from healthy women and from women with PMOS randomized to placebo or metformin, all with uncomplicated pregnancies. PMOS placentas showed transcriptional downregulation across multiple cell types and shifts in cell type proportions. Specifically, syncytiotrophoblasts exhibited reduced expression activity of growth hormone receptor signaling and glycosaminoglycan biosynthesis. Endothelial cells displayed diminished receptor tyrosine kinase pathway activity, including *VEGFC*, despite increased cell proportion and hypervascularity. Intercellular communication networks were globally suppressed, including reductions in PDGF signaling from Hofbauer cells to fibroblasts. Metformin induced limited molecular changes in PMOS placentas, with associated transcriptional shifts linked to lower birth weight and higher childhood BMI. Our findings revealed cell type–specific alterations in the PMOS placentas that showed limited responsiveness to metformin. This supports further exploration of mechanism-based therapeutic alternatives for future pregnancy management.

## Introduction

Polyendocrine metabolic ovarian syndrome (PMOS), previously termed polycystic ovary syndrome (Teede et al.), is the most common endocrine and metabolic disorder affecting 11-13% of women (Teede et al., 2023). It is associated with an increased risk of pregnancy complications, including miscarriage, gestational diabetes, and pre-eclampsia (Bahri Khomami et al., 2018, Bahri Khomami et al., 2024). Offspring of women with PMOS are often born with lower birth weight (Bahri Khomami et al., 2024) and experience early adiposity rebound with accelerated weight gain from puberty to adulthood (Koivuaho et al., 2019, Ollila et al., 2016). They also have a higher risk of developing cardiometabolic and reproductive disorders later in life (Yang et al., 2024), underscoring the long-term impact of PMOS. Growing evidence suggests that altered epigenetic and developmental programming due to hormonal dysregulation contributes to PMOS inheritance, as the genetic loci linked to PMOS account for only about 10% of its estimated 70% heritability (Stener-Victorin and Deng, 2021, Abbott et al., 2002).

The human placenta is recognized as a mediator of developmental programming and thus as a potential player associated with PMOS effects (Burton et al., 2016, Barker et al., 1990). It serves not only as an organ for nutrient and gas exchange but also as an active endocrine and metabolic regulator that profoundly influences both immediate pregnancy outcomes and long-term offspring health (Kramer et al., 2023, Gao et al., 2026). Placentas from women with PMOS show decreased volume and weight, with higher occurrences of inflammation, and villous immaturity, a histological pattern in which terminal and mature intermediate villi are under-represented relative to gestational age and the villous membrane retains a more immature architecture with fewer vasculo-syncytial membranes (Koster et al., 2015). These changes have been associated with genes like *FOS* and chemokines identified through RNA sequencing (Xie et al., 2024, Wang et al., 2020a). Therefore, improving placental function could potentially interrupt the cycle of disease transmission, positioning the placenta as both a key mediator and a therapeutic target (Stener-Victorin and Deng, 2024). However, the global transcriptional landscape of the placenta at cell-type resolution in the context of maternal PMOS remains uncharacterized and villous function depends on the coordinated activity of several distinct cell populations (Baergen et al., 2021, Derisoud et al., 2024, Liu et al., 2018, Vento-Tormo et al., 2018): The syncytiotrophoblast (STBs) forms the multinucleated interface which secretes hormone and allows nutrient and gas exchange; cytotrophoblasts constitute the progenitor pool that renews and fuses into this syncytium and therefore determines villous maturation; vascular endothelial cells (VECs) and perivascular cells (PVs) build and regulate the fetal capillary network that sets villous perfusion capacity; villous fibroblasts deposit and remodel the stromal extracellular matrix that supports villous architecture; and Hofbauer cells (HBCs), the resident fetal macrophages, contribute to immune surveillance, angiogenesis and stromal remodelling (Reyes and Golos, 2018).

Although PMOS is associated with increased risk of pregnancy complications, no specific interventions are recommended during pregnancy (Teede et al., 2023). Metformin, widely used for glycemic control in type 2 diabetes (Bailey and Turner, 1996) and prescribed off-label in non-pregnant women with PMOS, is often continued into pregnancies (Raperport et al., 2021). However, the randomized, placebo-controlled PregMet trials, in which metformin (2000 mg daily) or placebo was given from the first trimester (gestational week 5–12) until delivery, did not detect significant effect on gestational diabetes or preeclampsia (Fougner et al., 2008, Vanky et al., 2010b, Hanem et al., 2021). Follow-up studies raised concerns about offspring outcomes, including higher risk of overweight (Hanem et al., 2019b), and elevated circulating androgens in daughters (Hanem et al., 2021). The limited and inconsistent efficacy of current pharmacological interventions highlights the need to better understand placental molecular signatures in PMOS pregnancies. Key questions remain, such as which cell populations are affected, which transcriptional programs are disrupted, and which transcriptional differences are associated with metformin treatment. To address these questions, we applied single-nuclei RNA sequencing (snRNA-seq) to map the molecular landscape in term placentas from healthy controls and women with PMOS randomized to placebo or metformin during pregnancy in the PregMet trial (Vanky et al., 2010b, Lovvik et al., 2019). We focused on the placental villous tissue from uncomplicated pregnancies to reduce confounding effects of overt obstetric pathology. To add spatial and morphological context beyond single-nuclei transcriptomic analyses, we integrated H&E-based digital histology (HAPPY) (Vanea et al., 2024) and SpaTial Enhanced Resolution Omics-sequencing (Stereo-seq) (Zhao et al., 2025), enabling identification of specific histo-architectural alterations and spatially resolved gene expression. Together, these approaches provide a comprehensive, spatially informed framework for understanding placental cellular and molecular signatures in PMOS and evaluating metformin-associated alterations, with the goal of guiding future therapeutic interventions.

## Results

### Placental composition landscapes in polyendocrine metabolic ovarian syndrome with and without metformin treatment

To investigate the molecular basis of PMOS outcomes, we first performed bulk mRNA sequencing on placentas from 14 women with PMOS randomized to placebo (PMOSplac) and 16 women with PMOS randomized to metformin (PMOSmet) placentas from the PregMet1 trial (Vanky et al., 2010a). No clear separation between treatment groups were detected by principal component analysis (PCA), and no genes passed the significance threshold in or differential gene expression (DEG) analysis (Fig. S1A, Supplementary Table 1), consistent with the modest clinical effects reported for metformin on major pregnancy complications in this trial. As bulk transcriptomic profiling may mask cell-type–specific effects in heterogeneous tissue, we next selected a subset of these placentas, comprising PMOSmet (n=9), PMOSplac (n=9), and healthy Controls (Control, n=6), with similar maternal age, body-mass index (BMI), gestational age at delivery, delivery mode and birth weight (Supplementary Table 2). All placentas were selected from uncomplicated pregnancies to minimize secondary effects of pregnancy complications. Key hormonal measurements of testosterone (T), androstenedione (A4), sex hormone binding globulin (SHBG), and insulin level did not differ significantly between PMOSmet and PMOSplac across gestation (Mann-Whitney U test, Supplementary Table 2, Fig.1A). Bulk transcriptomic profiles of the selected samples remained indistinguishable by PCA between PMOSmet and PMOSplac, while both segregated from controls (Fig. S1B). Placental villous tissues were then profiled by snRNA-seq, H&E–based digital histology, and Stereo-seq spatial transcriptomics for downstream analyses (Supplementary Table 2, Fig. 1A).

**Fig. 1.**
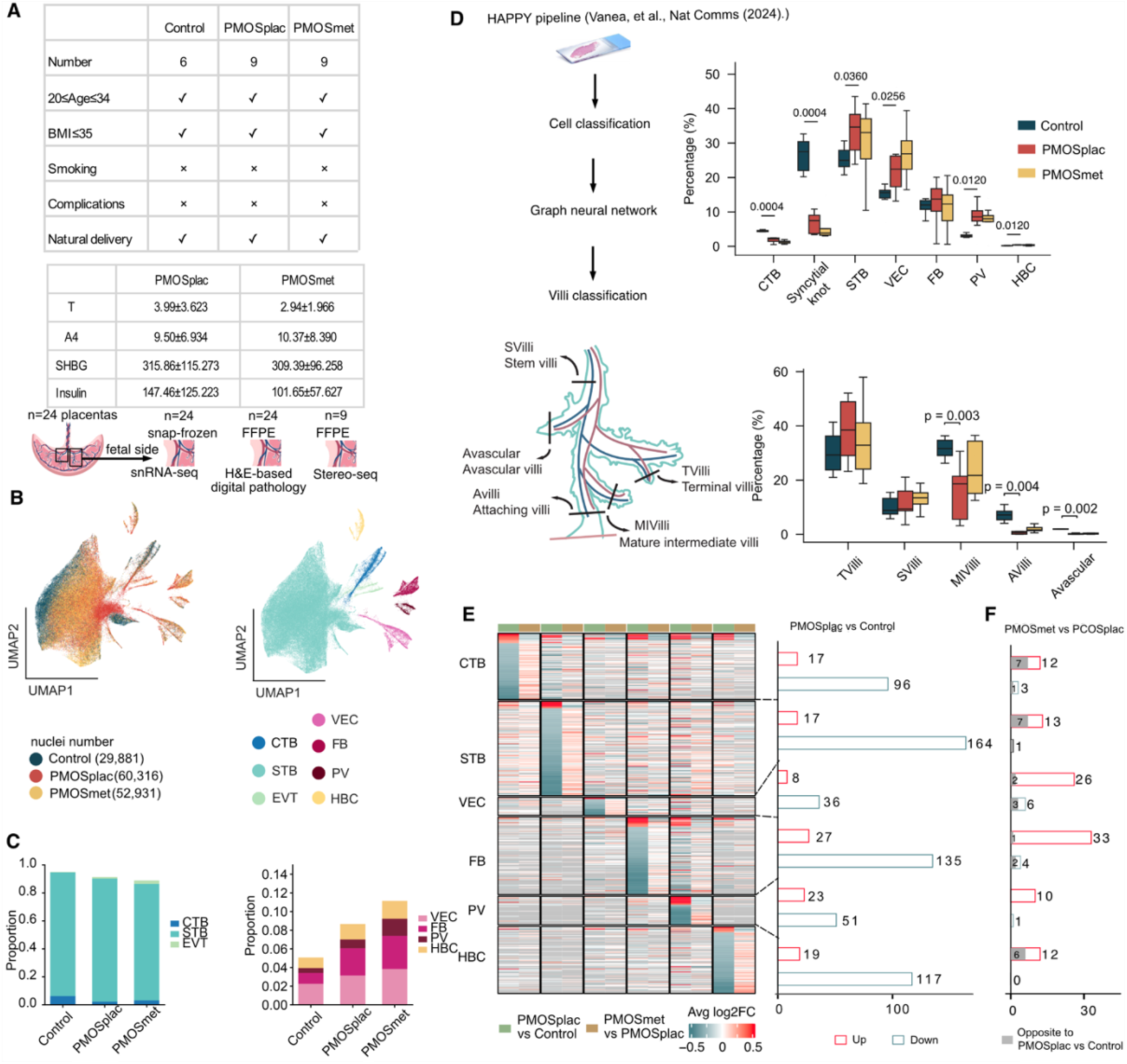
Placental composition and transcriptional profiling from women with polyendocrine metabolic ovarian syndrome (PMOS) with or without gestational metformin treatment. **(A)** Sample selection criteria, study design, and key hormonal measurements of testosterone (T), androstenedione (A4), sex hormone binding globulin (SHBG), and insulin level. **(B)** (left) UMAP embeddings of the single nuclei transcriptional profiles. (right) Cell type annotations of the nuclei clusters based on transcriptional profiles. **(C)** Cell type proportion captured by single nucleus RNA-seq across three groups. **(D)** Cell and tissue type proportions categorized by HAPPY^23^. Statistical significance was assessed using the Mann-Whitney U test (two-sided) for pairwise comparisons between Control (n=6) vs. PMOSplac (n=9) and PMOSplac (n=9) vs. PMOSmet (n=9) groups. Comparisons with p ≤ 0.05 are annotated on the figures. Data are shown as mean ± SD. **(E)** Average log2 fold change and number of DEGs comparing the placentas of PMOSplac with Control. **(F)** Number of DEGs between PMOSmet and PMOSplac. PMOSmet: PMOS randomized to metformin treatment group. PMOSplac: PMOS randomized to placebo group.

From snRNA-seq, we captured a total of 143,128 nuclei and identified seven placental cell populations: STBs, cytotrophoblasts (CTBs), extravillous trophoblasts (EVTs), fibroblast cells (FBs), PVs, VECs, and HBCs (Fig. 1B, Fig. S1C, Supplementary Table 3). EVTs were detected at a very low proportion of 1.3% (Fig. 1C), which is expected given that our sampling was primarily of placental villi excluding decidual tissue. PVs identified by high expression of contractile markers *ACTA2*, *TAGLN*, and *MYH11*, represented the rarest stromal population (1.1%), consistent with the sparse distribution of PVs around fetal vessels within villous stroma (Arutyunyan et al., 2023) (Fig. 1C). Compositional data analysis using scCODA (Buttner et al., 2021) indicated a reduction of STB and CTB proportions in both PMOS groups relative to Control (Fig. S1D, FDR<0.1). HAPPY, a hierarchical deep learning pipeline that identifies cell types and classifies tissue microstructures (Vanea et al., 2024), was applied to H&E sections from the same placentas. The histological composition showed that the reduction in trophoblast proportion was accompanied by fewer attaching villi and mature intermediary villi, a pattern consistent with villous immaturity (Koster et al., 2015), and this pattern was not appreciably shifted in the PMOSmet group (Fig. 1D, Fig. S2, Supplementary Tables 4, 5, Mann-Whitney U test).

### Transcriptomic differences associated with gestational metformin treatment

We examined how gestational metformin treatment related to PMOS-associated transcriptomic alterations (DEGs in PMOSplac versus Control). At the transcriptomics level, comparing PMOSplac to control, we identified 51 to 164 downregulated genes across major cell types in contrast to the number of upregulated genes, which ranged from 8 to 27 (Fig. 1E, Supplementary Table 6, 7). When comparing PMOSmet and PMOSplac, only a limited number of DEGs were detected between across cell types, most of which were higher in the metformin group (Fig. 1F). Among these DEGs, several overlapped with PMOS-associated signatures and were shifted toward control levels in the metformin group (Fig. 1F, Supplementary Table 6, 7). This finding further highlights the sensitivity of snRNA-seq in detecting subtle transcriptional differences.

In STBs, seven genes that were downregulated in PMOSplac showed expression closer to control levels in PMOSmet (Fig. 2A). Among these, *FBN2* encodes placensin, a glucogenic hormone that promotes trophoblast invasion and placentation (Yu et al., 2020), and serves as an extracellular matrix substrate for the protease ADAMTS6 in trophoblast niche maintenance (Wachter et al., 2022). *ELMO1* was likewise higher in PMOSmet, an established activator of Rac-dependent cell fusion in other syncytial tissues (Tran et al., 2022, Zang et al., 2024b). Notably, *ELMO1* expression is reduced in preeclamptic placentas (Zhang et al., 2024a), mirroring what we observed in PMOSplac. Expression *DPYD,* a rate-limiting enzyme in uracil or thymine catabolism (Nishimura and Naito, 2006) was also closer to control levels in PMOSmet. A similarly modest rescue effect was observed in other cell types, in which *PAPPA2* stood out. It is a protease that increases IGF bioavailability and regulates fetal growth and metabolism (Laursen et al., 2001). Increased *PAPPA2* in maternal blood is a strong predictor of severe preeclampsia (Macintire et al., 2014, Elovitz et al., 2025), and hypoxia-induced trophoblast stress (Wagner et al., 2011). Consistent with increased risk of preeclampsia in women with PMOS (Valdimarsdottir et al., 2024), *PAPPA2* expression was increased in CTBs, VECs and FBs in PMOSplac, whereas this difference was not reversed in the metformin group (Fig. 2A). Although our selected samples excluded cases with preeclampsia, the elevated *PAPPA2* in PMOSplac could indicate subclinical molecular dysregulation, and this difference was attenuated in the metformin group (Fig. 2A). Similarly, the upregulation of the aromatase *CYP19A1* in VEC, and of *ATG7* and *NRIP1* in HBC was not reversed by metformin (Fig. 2A). *ATG7* is a key autophagosome mediator implicated in placental homeostasis and previously identified as a PMOS biomarker (Lu et al., 2024b). *NRIP1* encodes a nuclear receptor coregulator for PPARs and estrogen receptors (Augereau et al., 2006).

**Fig. 2.**
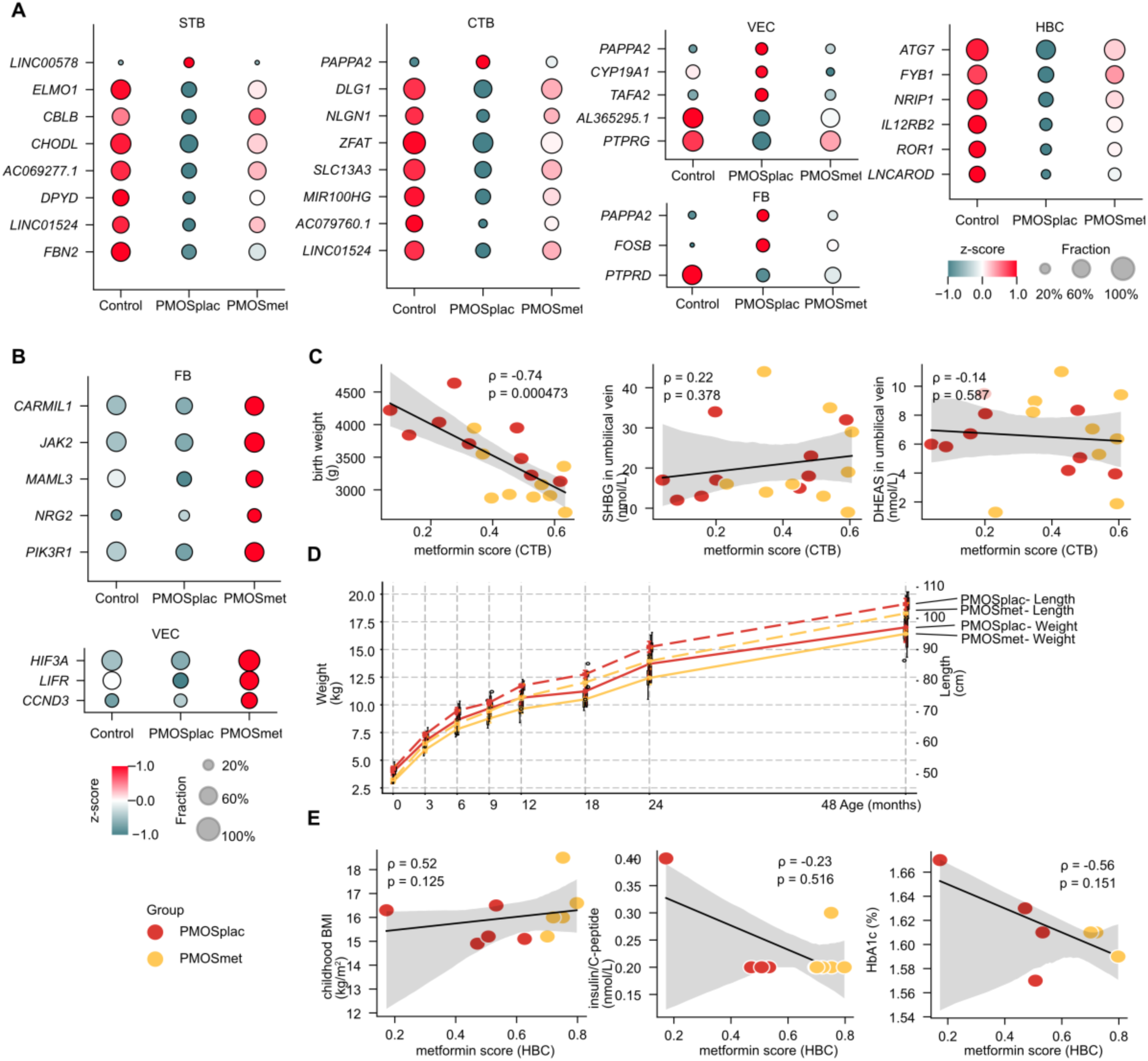
Gene expression affected by gestational metformin treatment of women with PMOS. **(A)** Differentially expressed genes (DEGs) between PMOSmet and PMOSplac. **(B)** DEGs in Fibroblasts and VECs that are induced by metformin but not as PMOS. **(C)** Correlation of metformin-associated transcriptional alteration score and birth weight, SHBG and DHEAS levels in umbilical vein. Correlations are Spearman’s ρ with two-sided P value and n = 9 placentas per group; The line is the linear regression fit and the shaded band is its 95% confidence interval. The gene set was the DEGs between PMOSmet and PMOSplac in CTB. And the enrichment was done by Gene Set Variation Analysis (GSVA). **(D)** growth curve of the offspring. The straight lines are body weight and use the y-axis to the left. The dashed lines are body length and use. The y-axis to the right. **(E)** Correlation of metformin-associated transcriptional alteration score and childhood body mass index (BMI), insulin/C-peptide and HbA1C levels. Correlations are Spearman’s ρ with two-sided P value and n = 5 placentas per group; The line is the linear regression fit and the shaded band is its 95% confidence interval. The gene set was the DEGs between PMOSmet and PMOSplac in Hofbauer cells. And the enrichment was done by GSVA.

We also identified 6 to 34 genes per cell type altered in PMOSmet placentas but not among PMOS-associated DEGs, potentially reflecting transcriptional responses associated with metformin unrelated to the PMOS signature (Supplementary Table 6). In fibroblasts, higher expression in PMOSmet was observed for the signaling and cytoskeletal regulators *PIK3R1*, *NRG2*, *MAML3*, *JAK2*, and *CARMIL1* (Fig. 2B), the latter linking cytoskeletal organization to inflammatory signaling and matrix remodeling (Wang et al., 2020b), suggesting increased fibroblast sensitivity to IL-1-mediated inflammatory signaling. In VECs, *HIF3A* and *LIFR* were higher in PMOSmet (Fig. 2B). *HIF3A* preferentially responds to hypoglycaemia rather than hypoxia (Heidbreder et al., 2007). LIF-LIFR signaling regulates cholesterol homeostasis through modulation of LDLR activity (Moran et al., 1997, Hemme et al., 2024).

We then calculated a metformin score measuring metformin-associated transcriptomic alterations using Gene Set Variation Analysis using DEGs from the PMOSmet versus PMOSplac comparison in each cell type (Hanem et al., 2019a). We correlated the offspring clinical phenotypes with the metformin score. Notably, the scores in CTBs correlated inversely with birth weight but showed no correlation with SHBG or dehydroepiandrosterone (DHEAS) levels in umbilical vein (Fig. 2C), in line with the clinical outcomes of metformin treatment in the PregMet cohort. For the metformin scores in other cell types, no correlation with birth weight or hormonal measurements was detected (Fig. S3A). The offspring born to PMOSmet mothers showed body weight catchup by 2 years-old (Hanem et al., 2018) (Fig. 2D). Moreover, the follow-up study of the children born to mothers in the PregMet cohort showed an increased BMI z-score in the PMOSmet group (Hanem et al., 2021). Interestingly, the metformin score in HBCs had the strongest correlation with childhood BMI among all cell types yet showed no correlation with insulin (C-peptide) or HbA1C levels (Fig. 2E), consistent with the reported lack of metabolic benefit of metformin in the cohort (Hanem et al., 2021). No other cell types showed any correlation with the childhood BMI or biochemistry measurements (Fig. S3B).

In summary, metformin was associated with only limited reversal of PMOS-associated transcriptional alterations, and a small number of additional gene expression changes largely distinct from the PMOS-associated signature. This aligns with clinical evidence of persistent pregnancy complications in PMOS and the absence of a clear obstetric benefit of metformin treatment (Vanky et al., 2010b, Hanem et al., 2019b, Vanky et al., 2004).

### Molecular signatures of syncytiotrophoblasts in polyendocrine metabolic ovarian syndrome

Given the limited transcriptional reversal of metformin, we next focused on cell type–specific PMOS-associated signatures as a possible foundation for placenta-targeted approaches in the future. As the predominant cell population in our dataset (123,617 nuclei, 86.4%), STBs were analyzed using Milo (Dann et al., 2022), an unsupervised, graph-based neighborhood analysis, to resolve PMOS-specific molecular signatures. STB nuclei were partitioned into 4,648 local neighborhoods (Fig. S4A, B). Differential-abundance testing (log₂FC > 2, spatial FDR < 0.05) identified 953 PMOS-enriched neighborhoods (30,194 nuclei) and 934 Control-enriched neighborhoods (20,973 nuclei), with the remaining neighborhoods showing transcriptional overlap (Fig. 3A and Fig. S4C). Restricting analysis to non-overlapping neighborhoods improved the signal-to-noise ratio and identified 786 downregulated and 53 upregulated genes in PMOS relative to controls (Fig. S4D, Supplementary Table 8).

**Fig. 3.**
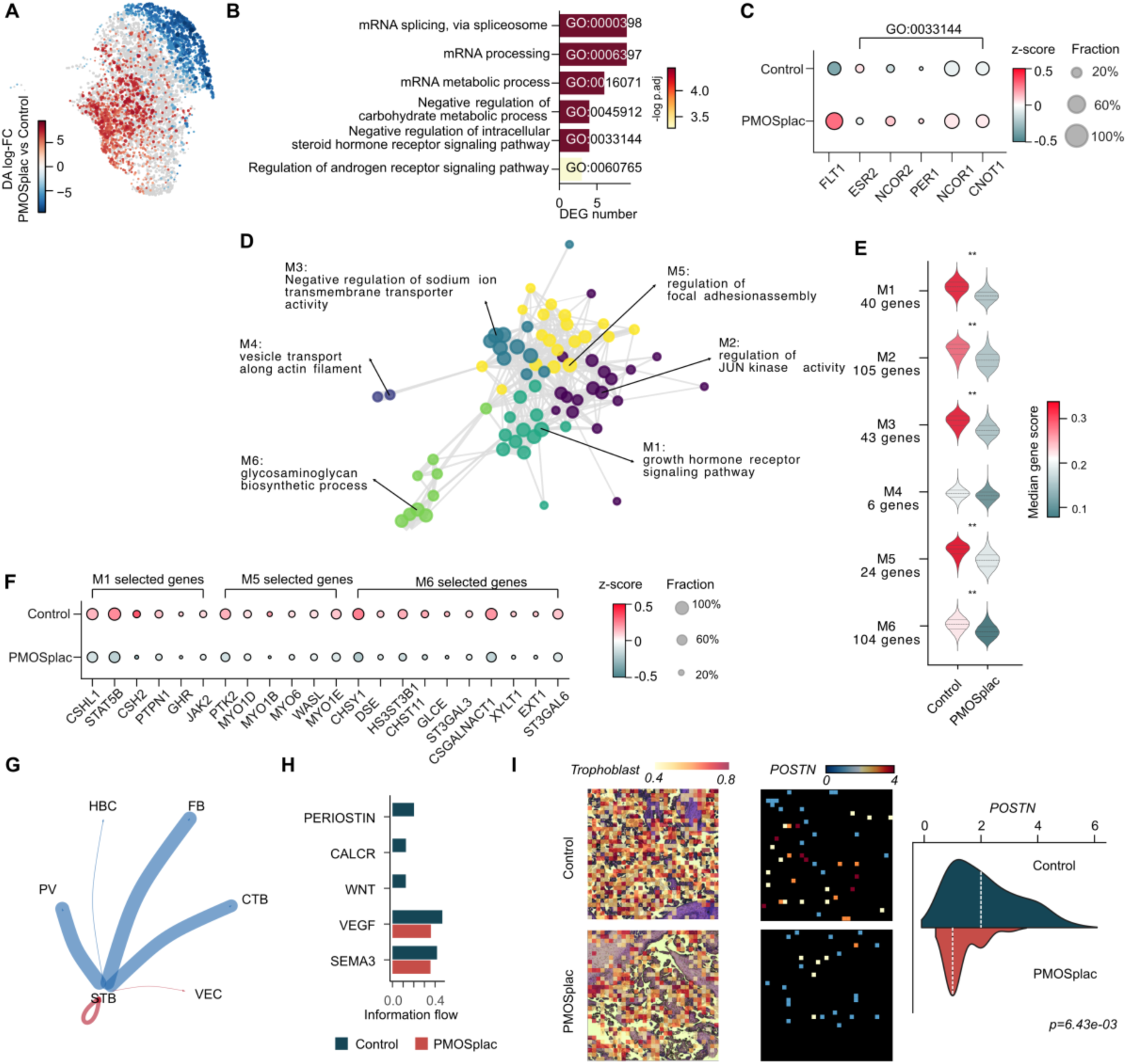
Molecular signatures of syncytiotrophoblasts (STBs) in placentas from women with PMOS. **(A)** Differential abundance test of STB nuclei neighborhoods between PMOSplac and Control. **(B)** Barplot showing the selected enriched gene ontology terms of the up-regulated genes in STBs of PMOSplac compared with Control. **(C)** Gene expression patterns of Estrogen Receptor and DEGs in “Negative regulation of intracellular steroid hormone receptor signaling pathway”. **(D)** Network of enriched GO terms from down-regulated genes in STBs of PMOSplac. **(E)** Gene set expression scores of genes in each GO community in Control-enriched nuclei neighbors, PMOSplac-enriched nuclei neighbors. ** indicates large effect size (d≥0.8), and *indicates medium effect size (0.5≤d<0.8), by Cohen’s d test. **(F)** Expression patterns of DEGs in GO pathways. **(G)** Differential communication strength between PMOSplac and Control to other cell types. Red arrows indicate increased while blue arrows indicate decreased communication probability in PMOSplac. Thicker line indicates greater changes. **(H)** Outgoing signaling pathways from STBs in PMOSplac and Control. **(I)** Magnified visualization showing (left) deconvoluted trophoblast proportion and *POSTN* (PERIOSTIN signaling) expression (middle) in regions of Control (top) and PMOSplac (bottom) samples. (right) ridge density plots comparing *POSTN* expression distributions between control and PMOSplac groups within the selected regions. Dashed lines indicate distribution of medians. Statistical significance was assessed by two-sided Wilcoxon rank-sum test. Each spatial bin represents a 50 × 50 µm area (bin100 resolution).

Among the 53 up-regulated genes, Gene ontology (GO) biological pathway analysis identified enrichment for mRNA splicing and processing (Fig. 3B). This is functionally relevant as STBs are terminally differentiated with limited transcriptional activity at term (Ellery et al., 2009), making post-transcriptional regulation a critical determinant of protein output in this compartment. Disrupted splicing is linked to preeclampsia and intrauterine growth restriction (Ruano et al., 2021) exemplified by excess sFLT1 (soluble VEGFR1) isoform driving endothelial dysfunction in preeclampsia (Turanov et al., 2018). Given that total *FLT1* transcript levels were higher in PMOSplac (Fig. 3C, Fig. S4E), altered processing may shift sFLT1 isoforms composition with pathological implications for angiogenesis. Upregulated genes were also enriched for negative regulation of intracellular steroid hormone receptor signaling (Fig. 3B), encoding transcriptional corepressors (*NCOR1, NCOR2*) and a coactivator *EP300*, which converge on nuclear steroid hormone receptors (Fig. 3C). Notably, *ESR2* (ERβ), was reduced in PMOSplac. Although STBs lack appreciable androgen receptor expression, testosterone-derived metabolites such as 3β-Diol can directly activate ERβ independent of androgen receptor (Jordan and DonCarlos, 2008), providing a plausible mechanism by which androgen excess in PMOS establishes negative feedback on ERβ signaling.

GO analysis of 786 downregulated genes identified 96 enriched terms (adjusted P-value <0.05, Supplementary Table 9), with top pathways including growth hormone receptor signaling, MAPK cascade, receptor tyrosine kinase signaling, and JAK-STAT activation (Fig. S4F). To reduce redundancy, we clustered the top 75% most significant GO terms by Jaccard similarity into six representative modules: M1 growth-hormone-receptor signaling pathway, M2 JUN-kinase activity, M3 negative regulation of sodium-ion transmembrane transporter activity, M4 vesicle transport along actin filaments, M5 focal-adhesion assembly, and M6 glycosaminoglycan (GAG) biosynthesis (Fig. 3D). Module activity scoring in STB nuclei confirmed significant downregulation of M1–M3 and M5–M6 in PMOS placentas, and a minor downregulation of M4 (Fig. 3E). Key molecular changes related to PMOS in M1 included downregulation of *CSH2* which encodes one isoform of placental lactogen (hPL) and the GHR– JAK2–STAT5B axis (Fig. 3F). This was consistent with reported reductions in growth hormone secretion in PMOS (de Boer et al., 2004). In M5, reduced *MYO1*, *MYO6*, and *WASL* expression were identified, implicating impaired actin-dependent membrane trafficking and receptor turnover in extensive STB syncytium in PMOS. In M6, suppression of GAG biosynthetic enzymes suggesting a similar pathological change as proteoglycan remodeling in preeclampsia (Heyer-Chauhan et al., 2014, Warda et al., 2008, Hassani Lahsinoui et al., 2021). Notably, heparin, a safe pharmacological GAG (Quenby et al., 2023, Zang et al., 2024a), has shown potential benefit in high-risk preeclampsia, and increases placental growth factor levels (Baroutis et al., 2025). This was relevant to observed fetal growth restriction in the offspring of PMOSplac (Talmo et al., 2024).

Intercellular communication was inferred using CellChat (Jin et al., 2021), which indicated that both the number and strength of predicted interactions were broadly reduced in PMOSplac (Fig. S4G–H). STB outgoing signaling was decreased towards most cell types, with only marginal increases toward VECs and STBs themselves (Fig. 3G). Pathway-level analysis identified several signaling pathways present exclusively in controls, including PERIOSTIN (Fig. 3H).

To further examine the spatial relationships between cell types, we performed Stereo-seq on three placentas per group with cell segmentation on the corresponding H&E images (Supplementary Table 10). We found that adequate transcript capture required bin100 resolution (>400 genes/bin), with lower resolutions yielding insufficient counts (Fig. S5A). Each bin100 entailed up to 10 cells (Fig. S5B). We then performed spatial deconvolution by mapping snRNA-seq nuclei to spatial bins using Tangram (Biancalani et al., 2021). The resulting spatial distributions were consistent with histological patterns, with trophoblast (STBs and CTBs) enrichment along the outer villous layer and vasculature (PVs and VECs) proportion concentrated around vessel structures (Fig S5C), supporting the deconvolution approach and enabling spatially resolved analysis of PMOS-associated transcriptional signatures. Spatial transcriptomics was consistent with reduced *POSTN* expression in PMOS, with a reduction in both the number of expressing bins and expression levels (Wilcoxon rank-sum test, p = 6.43 × 10⁻³) (Fig. 3I, Fig. S4I). POSTN activates WNT, TGF-β, and PI3K/Akt signaling through integrin binding, forming complex crosstalk (Wang et al., 2022), and has been proposed as a predictive marker for the success of endometrial implantation (Morelli et al., 2014, Eroglu et al., 2020), raising the possibility of a link to the increased odds ratio of miscarriage observed in women with PMOS.

### Reduced proliferative programs in cytotrophoblasts and vascular endothelial growth factor signaling in vascular endothelial cells

Compared with controls, CTBs from PMOSplac exhibited coordinated downregulation of canonical WNT and ERK signaling cascades (Fig. 4A, Supplementary Table 9), including key downstream effectors *LGR5*, *ERBB4* and *PDGFD*, and *MAP3K4* (Fig. 4B). WNT and ERK pathways crosstalk in trophoblasts and promote trophoblast proliferation and invasion during placental development (Meinhardt et al., 2016). This reduced expression in PMOS was consistent with impaired trophoblast growth and differentiation accompanied by the reduced CTBs and STBs proportions (Fig. S1D). Beyond proliferation, expression of transporters in CTBs were also compromised. Transporters mediating xenobiotic efflux *ABCB1* (Dunk et al., 2018), endocytic nutrient uptake *LRP2* (Marzolo and Farfán, 2011), metabolic substrate availability *SLC13A3* (Storm et al., 2016, Marzolo and Farfán, 2011), and Ca²⁺ homeostasis *SLC24A3* (Pajor, 2014) were downregulated in PMOS (Fig. 4B). Moreover, *SLC24A3* expression is reported to be regulated by the estrogen and hypoxia stress in placenta (Yang et al., 2013). Collectively, these expression changes are compatible with reduced efflux, nutrient supply and ionic homeostasis. Consistent with these expression changes, CTB-derived intercellular signaling towards other cell types was significantly reduced in PMOS (Fig. 4C). Specifically, PDGF, SEMA3 and BMP signaling were the most affected (Fig. 4D).

**Fig. 4.**
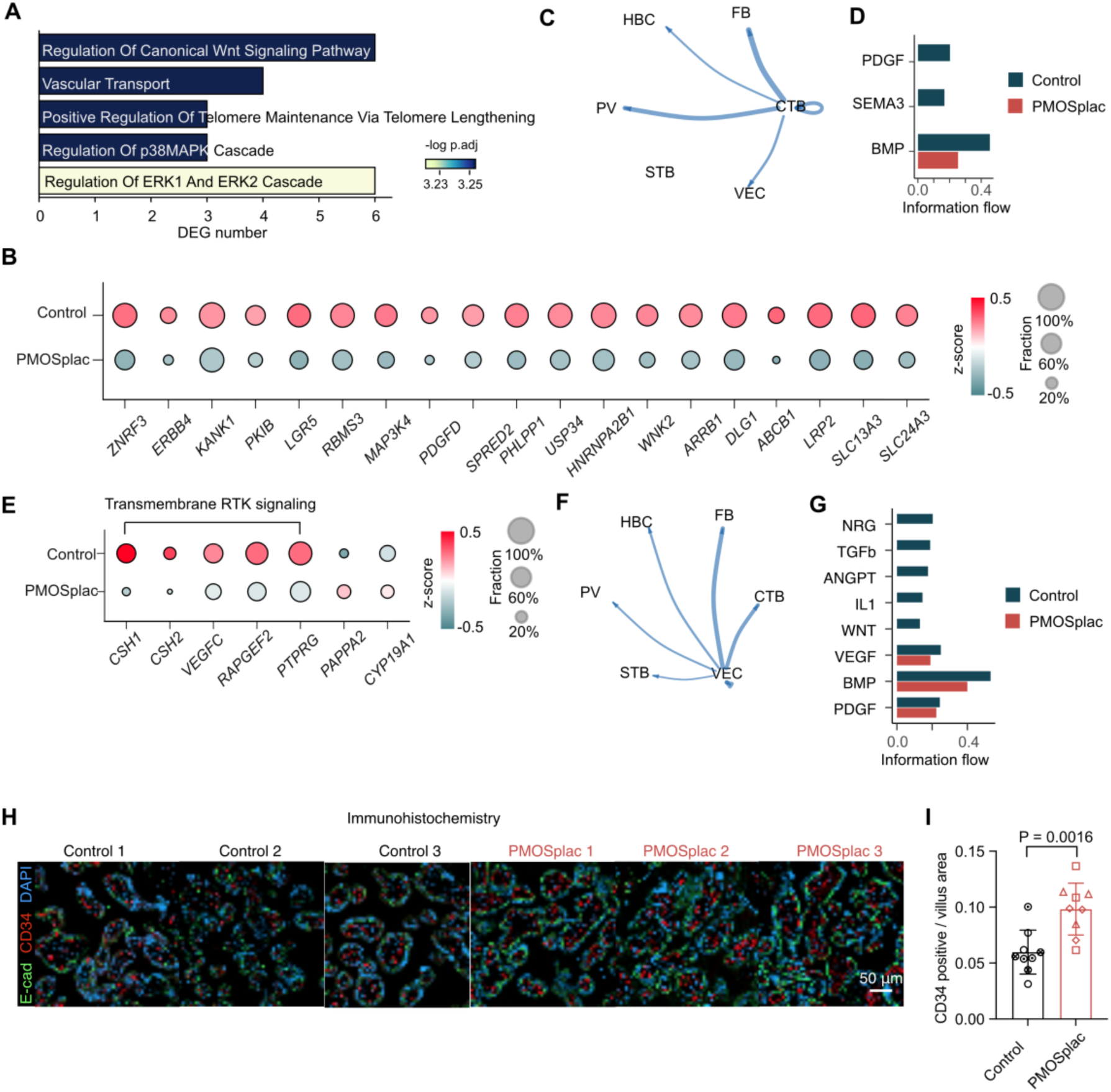
Transcriptional alterations of cytotrophoblasts (CTBs) and vascular endothelial cells (VECs) in placentas of women with PMOS. **(A)** The down-regulated GO terms in CTBs. **(B)** Dot plots showing the expression patterns of DEGs in CTBs between PMOSplac and Control, which involved in “Vascular transport”, “Regulation of canonical Wnt signaling pathway” and “Regulation of ERK1 and ERK2 cascade”. **(C)** Differential communication strength from CTBs between *PMOSplac* and Control to other cell types. Blue arrows indicate decreased communication probability in *PMOSplac*. Thicker line indicates greater changes. **(D)** Outgoing signaling pathways from CTBs in PMOSplac and Control. **(E)** Dot plots showing the expression patterns of DEGs in VECs between PMOSplac and Control, which involved in “Transmembrane Receptor Tyrosine Kinase signaling” and selected up-regulated genes. **(F)** Differential communication strength from VECs between PMOSplac and Control to other cell types. Blue lines indicate downregulated cell-cell communication in PMOSplac *vs* Control. Thicker line indicates greater changes. **(G)** Outgoing signaling pathways from VECs in PMOSplac and Control. **(H)** Immunohistochemistry images of the terminal villi. Red channel: CD34, Green channel: E-cad, Blue channel: DAPI. n=3 placentas per group. **(I)** Quantifications of placental vascular density. Villous regions were manually annotated based on E-cadherin signal in each image, and CD34 immunofluorescence was segmented using a pixel classifier in QuPath. Vascular density was expressed as the ratio of CD34-positive area to villous area. Three random fields were quantified as nested measurements within each placenta; group differences were tested using a nested two-tailed t-test. Data are shown as mean ± SD.

Interestingly, VECs showed downregulation of receptor tyrosine kinase signaling components, including *CSH1*, *CSH2* and *VEGFC* (Fig. 4E, Supplementary Table 9). Given the role of VEGFC in fetal lymphangiogenesis and erythropoiesis (Fang et al., 2016), its downregulation in VECs could indicate a reprogramming of vascular signaling, against lymphatic programs. In addition, VEC outgoing signaling towards other cell types was broadly reduced (Fig. 4F), with the most pronounced reductions in NRG and VEGF signaling pathways (Fig. 4G). NRG– ERBB4 signaling in VECs normally supports endothelial survival and angiogenesis under metabolic or inflammatory stress (Shakeri et al., 2018, Kalinowski et al., 2010). Moreover, terminal villi displayed increased vascular density shown by CD34 staining (Fig. 4H, I). The reduction of the signal alongside increased vascular density could reflect a compensatory regulatory response.

### Reduced extra cellular matrix pathways in fibroblasts and endocytosis pathways in Hofbauer cells

Villous FBs are the principal mesenchymal cells of the placental stroma responsible for extracellular matrix (ECM) deposition and structural support (Rossi et al., 2025). Using Jaccard similarity on pathway enrichment, we found that FBs exhibited coordinated downregulation of receptor tyrosine kinase signaling, ECM organization and positive regulation of cellular differentiation pathways in PMOS with signature genes highlighted in each pathway (Fig. 5A, Supplementary Table 9). Cell-cell communication analysis showed that signals from FBs to all cell types except for STBs were downregulated (Fig. 5B). The outgoing VEGF signaling from FBs to other cell types except HBCs were all increased, in synergy to the decrease of the outgoing VEGF signaling from STBs (Fig. S6A). However, inferred ligand *VEGFA* expression was not altered (Fig. S6B), suggesting this change could attribute to receptor expression in other cell types, such as *FLT1* in STB (Fig. 3C). Spatial deconvolution of Stereo-seq analyses confirmed FB-enriched bins distributed within the large villous stroma, where *PROS1* and *BMP5* exhibited modestly lower expression levels in PMOS compared with Control (Fig. 5D).

**Fig. 5.**
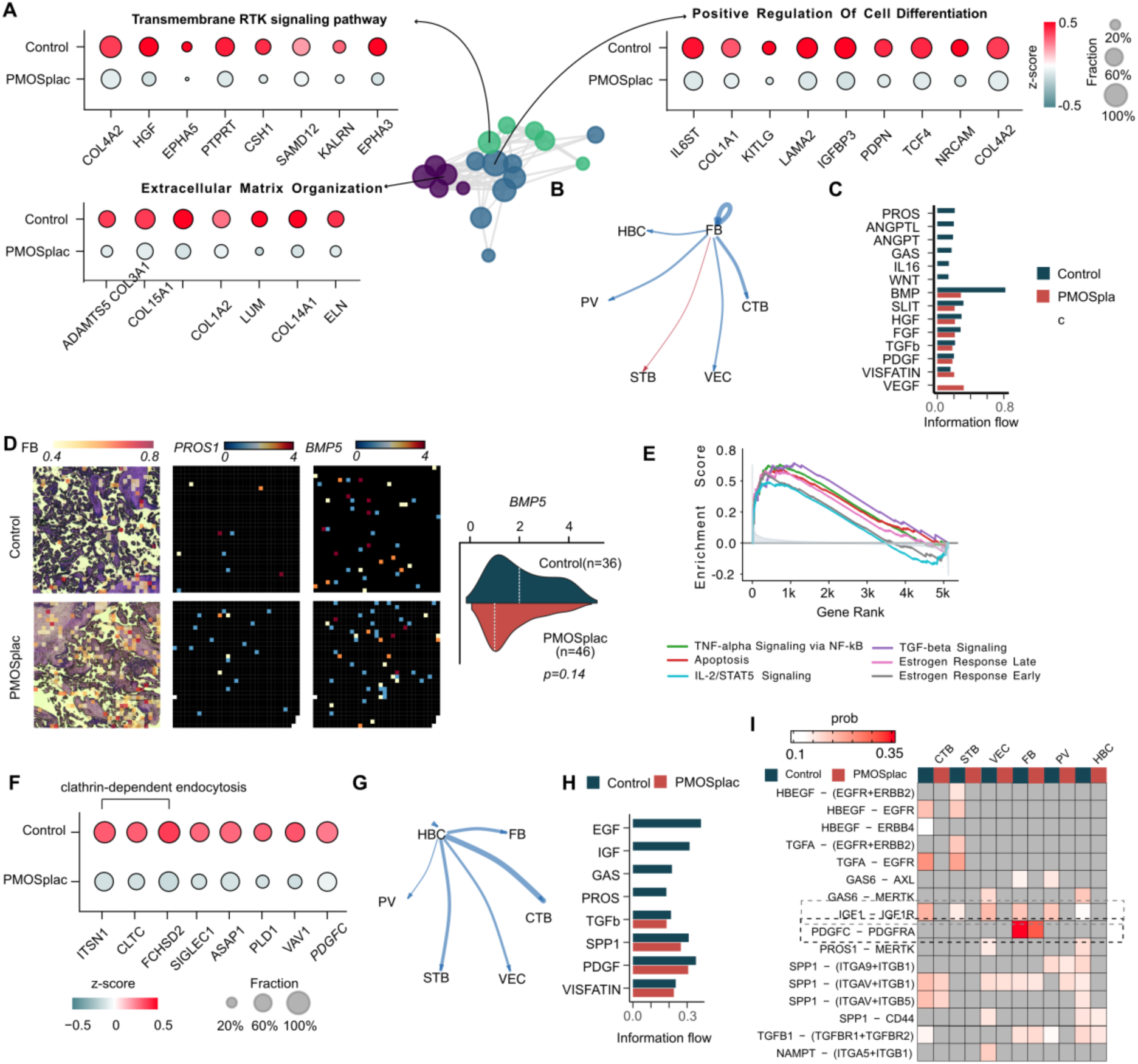
Fibroblasts (FBs) and Hofbauer cells (HBCs) expression signatures in placentas from women with PMOS. **(A)** Network of enriched GO terms from down-regulated genes in FBs of PMOSplac. and dot plots showing the expression patterns of DEGs involved in the selected GO terms. **(B)** Differential communication strength from FBs between PMOSplac and Control to other cell types. Red arrows indicate increased while blue arrows indicate decreased communication probability in PMOSplac. Thicker line indicates greater changes. **(C)** Outgoing signaling pathways from FBs in PMOSplac and Control. **(D)** Magnified visualization showing (left) deconvoluted FB proportion and *PROS1* (PROS signaling) and *BMP5* (BMP signaling) expression (middle) in regions of Control (top) and PMOSplac (bottom) samples. (right) ridge density plots comparing *BMP5* (BMP signaling) expression distributions between control and PMOSplac groups within the selected regions. Dashed lines indicate distribution of medians. Statistical significance was assessed by two-sided Wilcoxon rank-sum test. Each spatial bin represents a 50 × 50 µm area (bin100 resolution). **(E)** Enrichment analysis of gene expression HBCs between PMOSplac and Control. **(F)** Dot plots showing the expression patterns of DEGs involved in endocytosis and phagocytosis in HBCs. **(G)** Differential communication strength from HBCs between PMOSplac and Control to other cell types. **(H)** Outgoing signaling pathways from HBCs in PMOSplac and Control. **(I)** Dotplot showing the expression patterns of *PDGFC* (ligand) from HBCs and *PDGFRA* (receptor) from FBs.

HBCs showed no clear M1 polarization shift but displayed enrichment of inflammatory and stress-response pathways, including TNF-α, apoptosis, and IL-2/STAT signaling in PMOS compared with Controls (Fig. S6C, 5E). These gene expression patterns align with the hypothesis that tissue-specific immune responses underlie PMOS pathology, with a primed pro-inflammatory state similar to how macrophages are programmed in other tissues (Banerjee et al., 2023, De Robles et al., 2025).

Notably, GO analysis revealed suppression of “clathrin-dependent endocytosis” in HBCs in PMOS (Supplementary Table 9). The down-regulated genes include core vesicle formation and trafficking components such as *CLTC,* which encodes the clathrin heavy chain, *FCHSD2*, an adapter that recruits actin-polymerizing machinery to nascent pits (Xiao et al., 2018). And the downregulation of *ITSN1*, a scaffolds for clathrin, indicated impaired receptor internalization and vesicle budding (Jin et al., 2024) (Fig. 5F). Given the role of *ITSN1* in both endocytosis and early phagocytic events, its reduction also suggests compromised antigen uptake and signal integration (Jin et al., 2024). Additional decreases in *SIGLEC1* (CD169) and Fcγ receptor– associated genes *ASAP1*, *PLD1*, and *VAV1* further point to defective endocytic and immune-scavenging capacity (De Schryver et al., 2017, Brown et al., 1998, Liu et al., 2021, Bharti et al., 2007, Ali et al., 2013, Utomo et al., 2006) (Fig. 5F).

Most outgoing signals from HBCs were downregulated in PMOS (Fig. 5G). These included EGF, IGF, GAS, PROS and PDGF signals (Fig. 5H). Notably, PDGFC signaling is important for metabolic regulation and insulin sensitivity (Chen et al., 2020). It primarily targeted FBs and was diminished in PMOS (Fig. 5I). This reduce coincided with reduced ECM gene expression and receptor tyrosine kinase activity in fibroblasts (Fig. 5A). Consistently, *HGF* expression downstream of PDGF signaling in fibroblast was decreased in PMOS (Fig. 5A, Fig. S6D). Moreover, *FOXO1* was upregulated (Fig. S6D). It is a transcription factor negatively regulated by growth factor signaling including PDGF (Essaghir et al., 2009). Altogether, these observations are consistent with a shift toward a growth factor–poor stromal state.

### Coordinated signaling changes across placental cell types

To identify robust signaling alterations, we focused on pathways showing concordant reductions in both sender outgoing and receiver incoming signals, supported by transcriptional downregulation of the corresponding ligand or receptor (Fig. 6A, B, Supplementary Table 11). HBCs consistently showed reduced IGF signaling toward all villous cell types, with the most pronounced decrease toward CTBs. Reduced GAS6 signaling was found from HBCs to VECs (Fig. 6A). Interestingly, GAS6 was shown to regulate vascular homeostasis (Burstyn-Cohen and Fresia, 2023). Moreover, PROS1 signaling was reduced from FBs to HBCs and VECs. PROS1 contributes to endothelial barrier function, its reduced signaling could indicate compromised permeability regulation (Joussaume et al., 2024). POSTN signaling was reduced from STB to CTBs (Fig. 6A, 5B). And this could contribute to the suppressed WNT and ERK signaling activity observed in CTBs, as POSTN can induce WNT receptor engagement and MAP kinase pathway phosphorylation (Dorafshan et al., 2022).

**Fig. 6.**
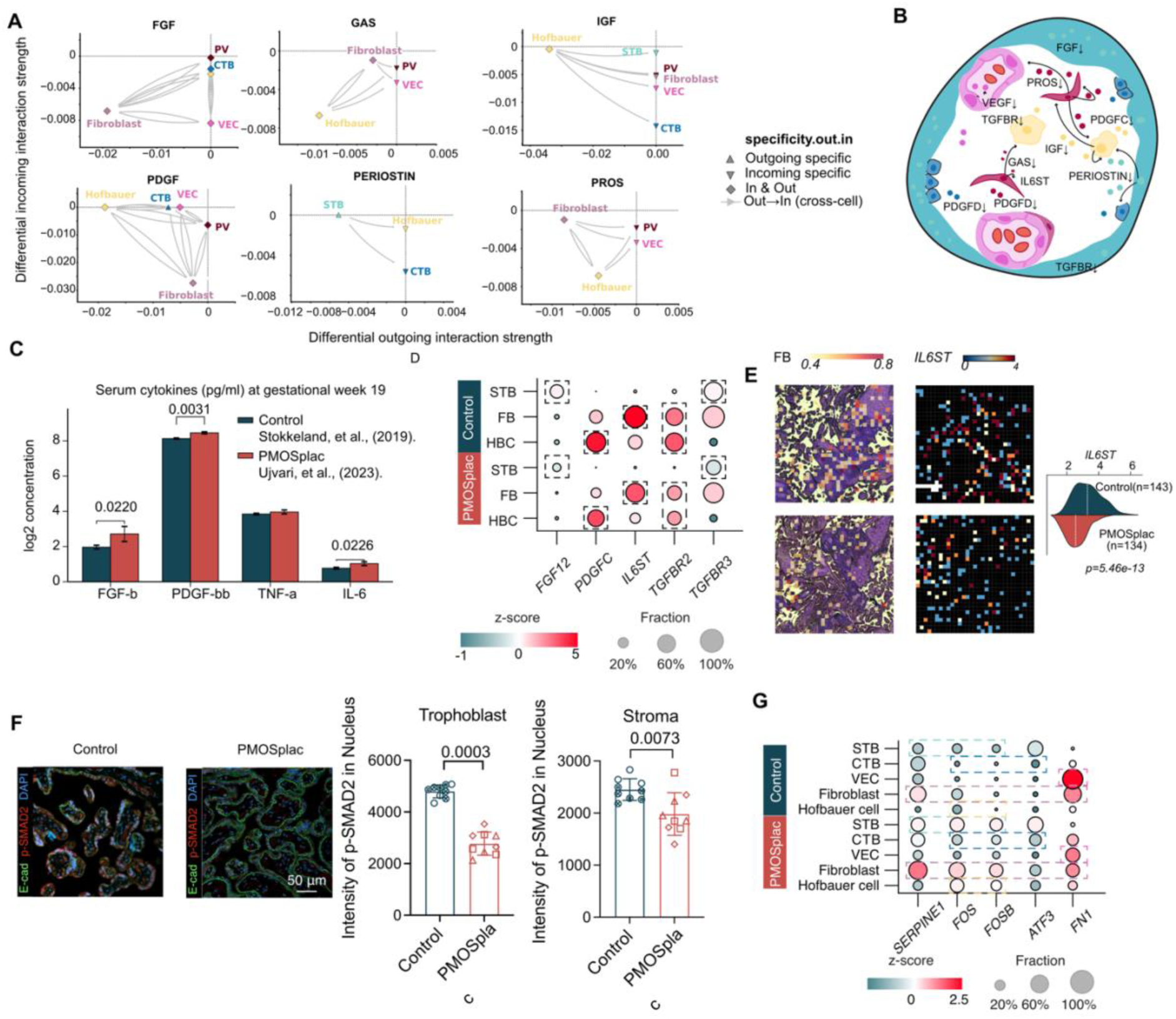
Altered intercellular signaling and serum cytokines in PMOS placenta. **(A)** Control-specific signaling inferred by CellChat and of which either ligand or receptor expression was differentially expressed. The arrows source from the outgoing specific cell type and target the incoming specific cell type. **(B)** Schematic overview of altered signaling networks within the placental villus microenvironment in PMOS. The diagram highlights key growth factors, cytokines, and receptors involved in cell-cell communication among placental cell types. downregulated arrows (↓) indicate expression levels of these specific factors in the PMOS condition. **(C)** Barplot comparing maternal serum cytokine levels (log2 concentration in pg/ml) at gestational week 19 between Control (dark blue) and PMOSplac (red) groups. Analyzed cytokines include FGF-b, PDGF-bb, TNF-α, and IL-6. Asterisks indicate statistically significant differences between the groups (Benjamin-Hochberg corrected p-value from Mann-Whitny U test). **(D)** Dot plot illustrating the comparative gene expression of selected signaling molecules and receptors across placental cell populations: Syncytiotrophoblast (STB), Fibroblasts (FB), and Hofbauer cells (HBC). The color bar corresponds to the expression z-score (red indicates higher relative expression; blue indicates lower), while the size of each dot represents the fraction of cells within that population expressing the gene (20% to 100%). Dashed boxes highlight genes differentially expressed in which cell types. **(E)** Magnified visualization showing (left) deconvoluted FB proportion and *IL6ST* expression (middle) in regions of Control (top) and PMOSplac (bottom) samples. (right) ridge density plots comparing and *IL6ST* expression distributions between control and PMOSplac groups within the selected regions. Dashed lines indicate distribution of medians. Statistical significance was assessed by two-sided Wilcoxon rank-sum test. Each spatial bin represents a 50 × 50 µm area (bin100 resolution). **(F)** Trophoblast and stromal regions were manually annotated based on E-cadherin signal. Nuclear mean fluorescence intensity of p-SMAD2 was quantified using DAPI to define nuclei in QuPath. Three random fields were quantified as nested measurements within each placenta; group differences were tested using a nested two-tailed t-test. Data are shown as mean ± SD. **(G)** Dot plot illustrating the comparative gene expression of selected signaling molecules and receptors across placental cell populations.

Next, we integrated these signaling with maternal circulating factors to decipher the potential receptor response to maternal factors. Targeted cytokine and growth-factor profiling of maternal serum at gestational week 19 showed higher circulating FGF-basic (FGF-B), PDGF-BB and IL-6 levels in women with PMOS compared with Control (Stokkeland et al., 2019, Ujvari et al., 2023) (Fig. 6C, Mann-Whitney U test). Yet placental expression of related signaling components *FGF12* in STBs and *PDGFC* in HBCs was reduced, suggesting compensatory response by the placenta (Fig. 6D). Despite elevated maternal IL-6 levels, *IL6ST* (gp130) was downregulated in FBs and also observed in Stereo-seq data, which is compatible with receptor desensitization mechanism (Fig. 6E). Moreover, downregulation of *TGFBR3* in STBs and *TGFBR2* in HBCs suggests attenuated TGF-β responsiveness (Fig. 6D). This was corroborated by reduced nuclear phospho-Smad2 immunofluorescence in both trophoblast and villous stromal cells (Fig. 6F).

Furthermore, *FOS* and *FOSB* were upregulated across multiple cell types, STB, CTBs, FBs, and HBCs (Fig 6G), and ATF3 was upregulated in CTBs and FBs. This was consistent with FOS/ATF3 identified as hub regulators in PMOS endometrium and ovary (Li et al., 2024). Furthermore, *FN1* was downregulated in VECs and FBs (Fig. 6G). It was previously identified as a hub protein reduced in placentas from women with PMOS (Sun et al., 2020). This is compatible with a shared disease-responsive transcriptional program across tissues.

## Discussion

Despite metformin being one of the most widely prescribed medications in PMOS pregnancy, randomized trials have not demonstrated improvements in their primary endpoints in women with PMOS (Vanky et al., 2010a, Vanky et al., 2004, Lovvik et al., 2019). And meta-analyses report only modest effects in early pregnancy (Devall et al., 2026, Zhao and He, 2022). Long-term follow-up of metformin-exposed offspring has even revealed increased childhood BMI (Hanem et al., 2019b). Why metformin shows limit clinical benefit, and whether it carries developmental risk, remains unresolved. One potential reason could be the placenta, which has not been examined at cellular resolution in women with PMOS. Here, by integrating analyses of snRNA-seq, spatial transcriptomics and digital histology on placentas from a randomized PregMet trial of metformin treatment throughout gestation in women with PMOS, we provide the first cell type-resolved transcriptomic map of placenta in this condition (Vanky et al., 2010a). Our findings identify latent molecular signatures that overlap with those reported in preeclampsia, even in clinically uncomplicated PMOS pregnancies (McGinnis et al., 2017, Nishizawa et al., 2008). Notably, the increased villous vasculature could reflect a compensatory response to the reduced outgoing signaling activity observed VECs. The limited clinical effects of metformin were mirrored at the molecular level, where only a small subset of PMOS-associated features altered by metformin and few additional transcriptional changes without significance.

Recently, several reference atlases of the healthy placenta have been published (Wang et al., 2026, Wang et al., 2024, Arutyunyan et al., 2023, Ounadjela et al., 2024). Our study extends these efforts into a disease context of PMOS pregnancies and further incorporates an intervention comparison. By focusing on clinically uncomplicated pregnancies of women with PMOS and offspring of normal birthweight, our design allowed identification of molecular alterations present in the absence of overt obstetric complications. Despite the absence of clinical complications, PMOS placentas exhibited coordinated molecular alterations across all major cell types. Previous studies have reported reduced placental size, villous immaturity, inflammatory infiltration, and altered nutrient transport and growth-factor signaling in PMOS (Koster et al., 2015, Tal et al., 2014, Jansson and Powell, 2013). Our multi-modal analyses are consistent with these observations and further resolved them to cell type-specific expression differences. This includes impaired progenitor renewal in CTBs, reduced intracellular transport and membrane integrity, and decreased receptor tyrosine kinase signaling in STBs. Moreover, previously reported hub regulators identified in other reproductive tissues in PMOS, including *FOS*, and *ATF3,* were also captured in our data. Thereby, we anchors these PMOS-associated disease molecules in the placenta for the first time.

Across cell types, reduced ligand availability converged on key growth factor pathways, including PDGF from HBCs, *CSH2* (encoding an hPL isoform) from STBs, and VEGF signaling from VECs. Reduced hPL trajectories have previously been associated with impaired fetal growth and lower birth weight (Abd El Aal et al., 2005). Strikingly, transcriptional signatures and impaired placental structural maturation linked to placental insufficiency and preeclampsia risk were detectable even in the absence of clinical complications. Elevated *FLT1* expression in STBs is a possible link to correspondence with the preeclampsia risk observed in PMOS pregnancies (Dasanu et al., 2011, Bodova et al., 2011, Teede et al., 2023). And the placental sFLT1-driven antiangiogenic profile that precedes overt preeclampsia (Zhang et al., 2024a). Reduction of *POSTN* has been reported as a predictive marker for endometrial implantation failure, and detected in amniotic fluid of pregnancies complicated by fetal congenital heart disease (Xia et al., 2023, Morelli et al., 2014, Eroglu et al., 2020). This warrants further investigation into its role in the potential abnormal fetal cardiac developmental in PMOS pregnancies. The collective signature raises the possibility that molecular reprogramming is detectable without clinical manifestations. Thus, stratification and intervention are speculated to be possible without overt obstetric complications and adverse offspring outcomes.

A striking feature of this latent phenotype is the presence of coordinated compensatory responses that appear to constrain progression to overt disease. The most prominent is a reorganization of placental VEGF signaling. Maternal serum VEGF is elevated in PMOS (Abd El Aal et al., 2005). Yet rather than amplifying this stress, the syncytial interface buffers it: STBs shift from VEGF sender to *FLT1*+ receiver, and FBs contribute a larger share of VEGF signaling to VECs. The elevated STB *FLT1* expression consistent with local buffering of excess maternal VEGF. This rewiring is structurally mirrored by increased villous vascular density, which we interpret as compensatory perfusion supporting fetal supply despite reduced ligand availability from growth factors elsewhere in the villous. We propose that the subclinical, uncomplicated clinical state of these PMOS pregnancies reflects successful but fragile placental adaptation, consistent with the allostatic load model, in which the physiological mechanisms the placenta normally uses to maintain homeostasis under stress become maladaptive when chronically or extremely activated, resulting in allostatic overload that contributes to disease (Rittenhouse et al., 2025, Premji et al., 2022, Premji et al., 2025). In our data, the molecules that may buffer maternal pro-angiogenic stress in this cohort overlap with those whose dysregulation has been associated with overt preeclampsia in other studies. This would reframe PMOS placental pathology not as static dysfunction but as a dynamic equilibrium between insult and compensation. When this equilibrium is tipped, it manifests as the 2-fold preeclampsia risk observed epidemiologically (Qin et al., 2013).

Against the PMOS-associated responses, metformin produced a narrow and selective transcriptional footprint. This includes *PAPPA2*, which is a strong predictor of severe preeclampsia, and *ELMO1*, a transcriptional hub in preeclamptic placentas (Zhang et al., 2024b, Sorensen et al., 2000). The core PMOS-associated responses were largely unchanged by metformin, providing a potential molecular explanation for its limited effects on primary clinical endpoints in trials of PMOS, gestational diabetes, type 2 diabetes, and maternal obesity trials (Feig et al., 2016, Morelli et al., 2014, Eroglu et al., 2020, Feig et al., 2020). Metformin-associated transcriptional changes in CTB correlated with lower birth weight, while corresponding changes in HBC showed a weaker correlation with childhood BMI. In FBs and VECs, metformin was associated with additional changes distinct from the PMOS signature, involving pathways central to compensatory VEGF signaling. Specifically, fibrosis regulators *JAK2* and *PIK3R1* in FBs; and *LIFR* and *HIF3A* in VEC were altered. One possibility is that these effects intersect with the compensatory responses that maintain the PMOS placenta in subclinical state. These molecular patterns are consistent with follow-up studies from PregMet trials and studies in gestational diabetes, showing increased central adiposity, elevated androgen levels, and altered metabolic profiles in metformin-exposed children(Chiswick et al., 2015, Hanem et al., 2019a, Hanem et al., 2021). Collectively, these suggest caution about dose escalation in the absence of a better mechanistic understanding.

Our findings carry three translational implications. First, current pharmacological strategies that target maternal systematic metabolism appear to have limited impact on PMOS-associated placental dysfunction and may intersect with the compensatory responses that accompany subclinical states (Brand et al., 2025). Second, the signatures we identify, particularly the growth factor, angiogenic pathways define candidate intervention nodes that are placentally tractable and avoid the constraints of systemic androgen blockade (Matsushita et al., 2018, Lu et al., 2024a). Third, the observation that clinically uncomplicated PMOS pregnancies may harbor a latent placental phenotype would support evaluating placental-function surveillance in PMOS pregnancy even when overt risk factors are absent. For example, sFLT1 monitoring could be a clinically available readout of compensation and may identify pregnancies in which compensation is failing before complications manifest.

Our findings should be interpreted in light of several limitations. The sample size, while sufficient for cell-type-resolved discovery, limits statistical power for subgroup stratification within PMOS and for detecting subtle treatment-associated differences. Placentas were sampled at delivery. Therefore, the temporal trajectory by which the compensatory equilibrium emerges will require longitudinal sampling and first-trimester biopsy approaches. The correlation between placental transcriptional alterations and offspring outcomes should be considered hypothesis-generating. Targeted follow-up in matched maternal–placental– neonatal triads will be required to substantiate the exposure–programming relationships. Finally, transcriptional changes do not directly index protein abundance or signaling activity. Functional validation of priority compensatory (*FLT1*, *VEGF*) and disease (*SERPINE1*, *PAPPA2*, *LIFR*) expression in maternal serum or umbilical blood is the next mechanistic step.

Nevertheless, the strength of our study is to provide disease state data that examine maternal pathophysiology, that will drive progression from descriptive mapping of healthy placental development to actionable target identification in pathologies. By studying placentas from clinically uncomplicated PMOS pregnancies without overt dysfunction or obstetric complications, we describe a state marked by molecular signs of adaptation. We show that metformin treatment was associated with a small number of additional transcriptional changes, while core disease signatures remain largely unchanged. Future clinical trials of pharmacological interventions during pregnancy should therefore incorporate concurrent placental profiling to evaluate both therapeutic efficacy and off-target placental responses.

## Methods

### Sample collection and RNA sequencing

Placental samples were collected from PregMet1 study (Vanky et al., 2010a). Sample transfer and research from PregMet1 was approved by Etikprövningmyndigheten (2022-04167-01). Only placentas from uncomplicated term pregnencies (≥ 37 gestational weeks), with spontaneous conception, vaginal delivery were selected. Within the PregMet1 trial, women were randomized to metformin (2000 mg daily) or placebo from the first trimester until delivery; both randomized arms were sampled with identical criteria. Pregnancies complicated by gestational diabetes mellitus, pre-eclampsia, other hypertensive disorders of pregnancy, or fetal growth restriction, as well as pregnancies with reported smoking, alcohol or drug use, and women with pre-existing diabetes at study entry were excluded.

The placental samples were collected shortly after delivery of the placenta. Villous parenchyma biopsies of 4–5 mm width were excised from the fetal side of the placenta, within approximately 2-4 cm of the umbilical cord insertion. Chorionic plate, the basal plate and any adherent decidua were deliberately excluded. Overtly infarcted, calcified or haemorrhagic areas were avoided. Biopsies were rinsed briefly to remove maternal blood and snap-frozen in liquid nitrogen.

Control samples were collected in collaboration with Karolinska University Hospital. This sample collection was approved by Etikprövningmyndigheten (2022-03542-01, 2022-06643-02, 2023-02270-02). Placentas were collected and processed immediately after delivery. During the whole procedure, samples were kept on ice. Sampling was performed close to the umbilical cord, following the same anatomical criteria described above for the PregMet1 biopsies. A full thickness sample was fixed with 4% PFA and stored at a 4℃ fridge before further processing for histological analysis. For transcriptome analysis (bulk and single nuclei RNA-seq), placental samples were snap-frozen in liquid nitrogen and stored at 80°C for later use. Control placentas were obtained from women with normal, uncomplicated singleton pregnancies, defined as spontaneous conception, no diagnosis of PMOS, no gestational diabetes mellitus, no hypertensive disorder of pregnancy including pre-eclampsia, no fetal growth restriction or macrosomia, no known fetal anomaly, delivery at term (≥ 37 completed gestational weeks) with an appropriate-for-gestational-age birthweight, and no reported smoking, alcohol or drug use during pregnancy, no pre-existing diabetes and no chronic medication known to affect placental function.

#### Single nuclei isolation

Nucleus isolation protocol was adapted from a method optimized for human placental tissue by Ludivine Doridot and colleagues (Institut Chochin, France), as described previously (Jiang et al., 2026). Briefly, approximately 30mg of frozen placental tissue was finely chopped on dry ice and transferred to a Douncer homogenizer containing cold NP-40 lysis buffer supplemented with RNAse inhibitor. Following a 10-minute incubation on ice, wash buffer (cold PBS containing 2% BSA and RNAse inhibitor) was added and homogenization was performed using 10 strokes with the loose pestle. The homogenate was sequentially filtered through 100 µm, 40 µm and 20 µm mesh filters. Nuclei concentration was determined using trypan blue staining and an automated cell counter (Bio-Rad TC20), and nuclei quality was assessed by microscopic visualization (EVOS XL Core, Thermo Fisher Scientific).

#### 10x Genomics Library Preparation and Sequencing

High-quality nuclei suspensions were processed according to manufacturer’s instructions for the Chromium Single Cell 3’ Kit v3.1 (10x Genomics). Library preparation was performed to obtain 10000 nuclei per reaction. Libraries were sequence to a depth of approximately 90 GB per sample (pair-end, 150bp), on an Illumina NovaSeq 6000 platform (Novogene, United Kingdom)

#### snRNA-seq data quality control, low-dimensional embedding, clustering and annotation

Gene counts were obtained by aligning reads to the GRCh38 genome using Cell Ranger software (10x Genomics). Cellbender (Fleming et al., 2023) was used to remove background noise. The quality control, data normalization, feature selection, low-dimensional embedding, and clustering were performed using scanpy v1.11.1 (Wolf et al., 2018). Specifically, using Scrublet (Wolock et al., 2019), cells that were labeled as doublets were removed. The principal components were decomposed from top 3,000 highly variable gene expression matrix. We selected the set of principal components relevant based on elbow plots as the space for neighbors finding. High resolution nuclei clusters were identified using the Leiden clustering algorithm. The nuclei data were projected on the uniform manifold for visualization. We annotated cell types using previously published marker genes and single-cell RNA-sequencing data (Derisoud et al., 2024).

#### Differential Abundance Test

The compositional changes across all cell types were performed using scCODA v0.1.9 (Buttner et al., 2021), which is a Bayesian model for compositional single-cell data analysis. The cutoff value for “false discovery rate” was set as 0.1. “HBCs” had the lowest dispersion and thus was set as the reference cell type.

The compositional changes within single cell type were tested using Milo in pertpy v1.0.0 (Heumos et al., 2026). Specifically, it assigned cells to neighborhoods on the k-nearest neighbors graph (k-NNG). Representation of the cells on 30 PCs was used for building the k-NNG. The differential abundance of the neighborhoods between the Control and the PMOSplac was then computed. The neighborhoods were grouped into Control-enriched, PMOSplac-enriched, and mixed. The control nuclei in the Control-enriched neighbors, and the PMOSplac nuclei in the PMOSplac-enriched neighbors were considered as Control specific and PMOS specific nuclei and was retrieved for DEGs.

#### Differential expression analysis

We performed differential expression analyses using MAST v1.32.0 “∼ 1 + group + sex” on log-normalized counts. Such analysis is deployed in R Seurat package v5.3.0 FindMarkers function, with false discovery rate (FDR) was estimated with Benjamini-Hochberg method (Butler et al., 2018). Genes with more than one count in at least 1% of cell, absolute differences in percentage of expression over 10%, average log2-fold change > 0.25 and FDR < 0.05 were considered.

Gene Ontology enrichment analyses were performed using GSEApy v1.1.8. Specifically, Fisher’s exact test was used to test the overrepresentation of the gene sets of interest from this study in known gene sets from MsigDB Hallmark, KEGG, and GO Biological Process databases. The p-value from the test was adjusted with the Benjamini-Hochberg method. The metformin score, that is, the metformin-associated transcriptional alteration score, was computed per sample and per cell type as follows. For each cell type, counts were first aggregated across nuclei of that cell type within each placenta to give one pseudo-bulk profile per sample, and pseudo-bulk profiles were library-size normalised and log-transformed. For each cell type, the gene set used for scoring was defined as the set of DEGs identified in the PMOSmet versus PMOSplac comparison for that same cell type, using the thresholds described above; up- and down-regulated genes were combined into a single set, and cell types with fewer than five DEGs were not scored. Enrichment of this gene set in each pseudo-bulk sample was then quantified using Gene Set Variation with default parameters for the Gaussian kernel appropriate to log-transformed continuous data, yielding one score per sample per cell type (Hanzelmann et al., 2013). Scores were computed across all three groups so that Control samples could be positioned relative to the treatment contrast, and were then related to offspring clinical variables (birth weight, umbilical vein SHBG and DHEAS, childhood BMI, C-peptide and HbA1c) using two-sided Spearman correlation, with the sample sizes stated in the corresponding figure legends.

#### Cell-cell communication inference

Cell-cell interactions were investigated using CellChat v2.2.0 (Jin et al., 2021). The interaction in each group was considered valid with “trimean” and filtered if expressed in less than 10 nuclei. Cell type level difference in the outgoing strength difference between groups (simply group A-group B) was calculated and visualized. Differences in ligand-receptor pairs were determined using “netVisual_bubble” function comparison parameter and considered significant with a *P* value of <0.05. CellChat infers signalling from the coordinated expression of ligand-receptor pairs and does not measure protein interaction directly, so all reported communication changes are inferential. For the syncytiotrophoblast, which is a single multinucleated syncytium rather than a population of separate cells, the inferred STB-to-STB signalling represents co-expression of ligand and receptor across STB nuclei within one continuous cytoplasmic compartment and is interpreted as autocrine or intra-syncytial signalling potential rather than as communication between distinct cells.

#### Maternal circulating factors

Maternal serum was collected at gestational week 19 and analysed by targeted multiplex cytokine and growth-factor profiling, including FGF-basic, PDGF-BB, TNF-alpha and IL- 6, as described previously (Stokkeland et al., 2019, Ujvari et al., 2023). Circulating factors were measured at a single gestational time point and in the subset of women for whom serum was available. Because the distributions were not assumed to be normal and group sizes were small, between-group comparisons (PMOSplac versus Control) were performed with the two-sided Mann-Whitney U test, and p values were adjusted across the analysed factors with the Benjamini-Hochberg procedure; adjusted p values below 0.05 were considered significant.

#### Predicting cell types and tissue types across human placenta H&E images

Histology slides were prepared using a standard H&E staining procedure. As per clinical guidelines, placental villi were sampled and 5 μm thickness slices were generated. Slides were scanned and digitized using Zeiss, a scanner at x20 magnification. Nuclei localization, cell classification, and tissue classification were performed using HAPPY pipeline (Vanea et al., 2024).

#### Immunofluorescence staining

Paraffin-embedded placental tissue sections (5 μm) were mounted on slides and dried overnight, followed by standard deparaffinization and antigen retrieval in Tris/EDTA buffer (pH 9.0) using microwave heating. Sections were then washed with PBST (PBS + 0.1% Tween-20), blocked with 2% donkey serum and 4% BSA for 30 min at room temperature, and incubated overnight at 4°C with primary antibodies diluted in PBST containing 1% donkey serum and 4% BSA. Primary antibodies used included anti-E-cad (R&D Systems, AF648), anti-CD34 (Atlas Antibodies, HPA036723), and anti-p-Smad2 (Millipore, 04-953). After washing, sections were incubated with secondary antibodies for 1 h at room temperature in the dark. Images were acquired using a Zeiss LSM 880 and analyzed with QuPath (v0.6.0).

#### Stereo-seq sample processing and sequencing

The sample library of Stereo-seq was constructed according to the manufacturer’s protocol for the Stereo-seq Transcriptomics N Kit V1.0 (MGI, 211KN114). Paraffin-embedded tissue sections (5 µm thick) were mounted onto the Stereo-seq Chip N Slide (1 cm × 1 cm), dried at 42 °C for 3 hours, and then baked overnight at 37 °C. The sections were subsequently deparaffinized, stained with hematoxylin and eosin, and imaged. The slides were then decolorized and de-crosslinked, followed by immersion of the tissue-mounted Stereo-seq Chip Slide in pre-cooled methanol for fixation at −20 °C for 20 minutes. Placental tissue sections attached to the chip were permeabilized at 37 °C for 30 minutes, followed by reverse transcription at 42 °C overnight. The cDNA released in situ was then collected, amplified, and purified. Barcodes were subsequently added, and rRNA was removed according to the manufacturer’s protocol for the SEQuoia RiboDepletion Kit (BIO-RAD, 17006487). Finally, DNA nanoballs (DNBs) were generated and loaded onto the flow cell for sequencing on the MGI DNBSEQ-G400RS platform.

#### Stereo-seq data analysis

Stereo-seq data were processed using the Stereo-seq analysis software suite (Gong et al., 2024). Specifically, the read files were processed using SAW v8 (https://github.com/STOmics/SAW). At the bin100 and single-nucleus resolution, we deconvoluted the cell types per bin100 based on segmentation using corresponding H&E images (0.447 um/pixel). To accurately estimate cell density per spatial spot, we performed nuclei segmentation on the histological images. High-resolution H&E images were normalized and segmented using a watershed algorithm implemented in StereoMap v<u>4.0 (</u>https://en.stomics.tech/resources/software-archives.html). We detected 65,208 to 203,969 segmented cells, with mean cell area of 112 µm^2^ per sample. The resulting segmentation shapes were used to calculate the number of nuclei per bin100 spatial using SpatialData.aggregate() in spatialdata v0.6.1 (Marconato et al., 2025). Bins containing fewer than one segmented cell were excluded from analysis.

A snRNA-seq reference for each group was prepared to guide the spatial deconvolution. To align the reference composition with the specific histological context of each sample, we applied a proportional downsampling strategy. We defined expected cell type proportions using the inferred proportion on the adjacent sections as processed in “*Predicting cell types and tissue types across human placenta* H&E *images”* section and subsampled the scRNA-seq reference to match these distributions, ensuring that rare cell types were sufficiently represented while mitigating compositional biases. Cell types with fewer than 10 cells were excluded. Feature selection was performed by identifying the top 100 marker genes per cell type using the Wilcoxon rank-sum test (scanpy.tl.rank_genes_groups) on log-normalized expression data. The union of these marker genes, intersected with genes detected in the spatial data, was used as the training gene set for learning the cell-to-spot mapping using Tangram (v 1.0.4) (Biancalani et al., 2021).

We used the default "cells" mapping mode, which learns a probabilistic assignment of individual single cells to spatial locations. The mapping was optimized over 1,000 epochs with a density prior derived from the aggregated cell counts per bin, normalized to a probability distribution, guiding the spatial allocation of cells proportionally to the observed cell density. Following the probabilistic mapping, cell type annotations were projected onto the spatial bins to derive per-cell-type enrichment scores. A per-bin proportion normalization was also computed, in which each spot’s cell type scores were divided by their sum so that values represent the estimated fractional cell type composition within each spot (summing to 1).

## Data Availability

The snRNA-seq data is available on Human Cell Atlas CELLXGENE Discover and spatial transcriptomics data is available on EMBL-EBI’s BioImage Archive. Code for snRNA-seq data quality control, data normalization, feature selection, low-dimensional embedding, and clustering were performed using scanpy is available at https://github.com/brainfo/ scutil. The spatial analysis pipeline, utilizing spatialdata and Tangram, is available at https://github.com/brainfo/ezsp. snRNA-seq data is available at human cell atlas: https://cellxgene.cziscience.com/collections/dbf29cf0-6f21-4b08-990d-5bfee327cd50 and spatial data is available in the BioStudies database (http://www.ebi.ac.uk/biostudies) under accession number S-BIAD3632.

## Author contributions

Q.D., E.S.-V, and H.J. conceived and designed the overall study. H.J. performed the data analyses of the RNA sequencing (RNA-seq), single nuclei RNA-sequencing (snRNA-seq), H&E imaging, and Stereo-seq. X.W. performed placental tissue section and H&E staining, Stereo-seq, and immunohistochemistry with the help of D.P. and P.R.J. E.V provided the placenta samples from PregMet cohort, and maternal serum, offspring metabolic measurements. E. D prepared libraries for RNA-seq and snRNA-seq. D.P, E.D, E.N, A.Z, S.S, M.H, C.M.L collected placenta from recruited mothers at Karolinska University Hospital. E.S.-V, and Q.D. provided conceptual and methodological suggestions and feedback. H.J. and Q.D. wrote the manuscript, to which all other authors provided input.

## Acknowledgements

This work was supported by the Swedish Medical Research Council. QD was supported by the Knut and Alice Wallenberg Foundation (2019.0211), Karolinska Institutet faculty funded position, KID grant, and Barndiabetesfonden. QD and E.S-V was supported by Diabetes foundation. HJ was supported by the Karolinska Institutet-China scholarship council program, (HJ). E.S.-V was supported by Distinguished Investigator Grant, Endocrinology and Metabolism from Novo Nordisk Foundation (NNF22OC0072904)

---

**Fig. S1.**
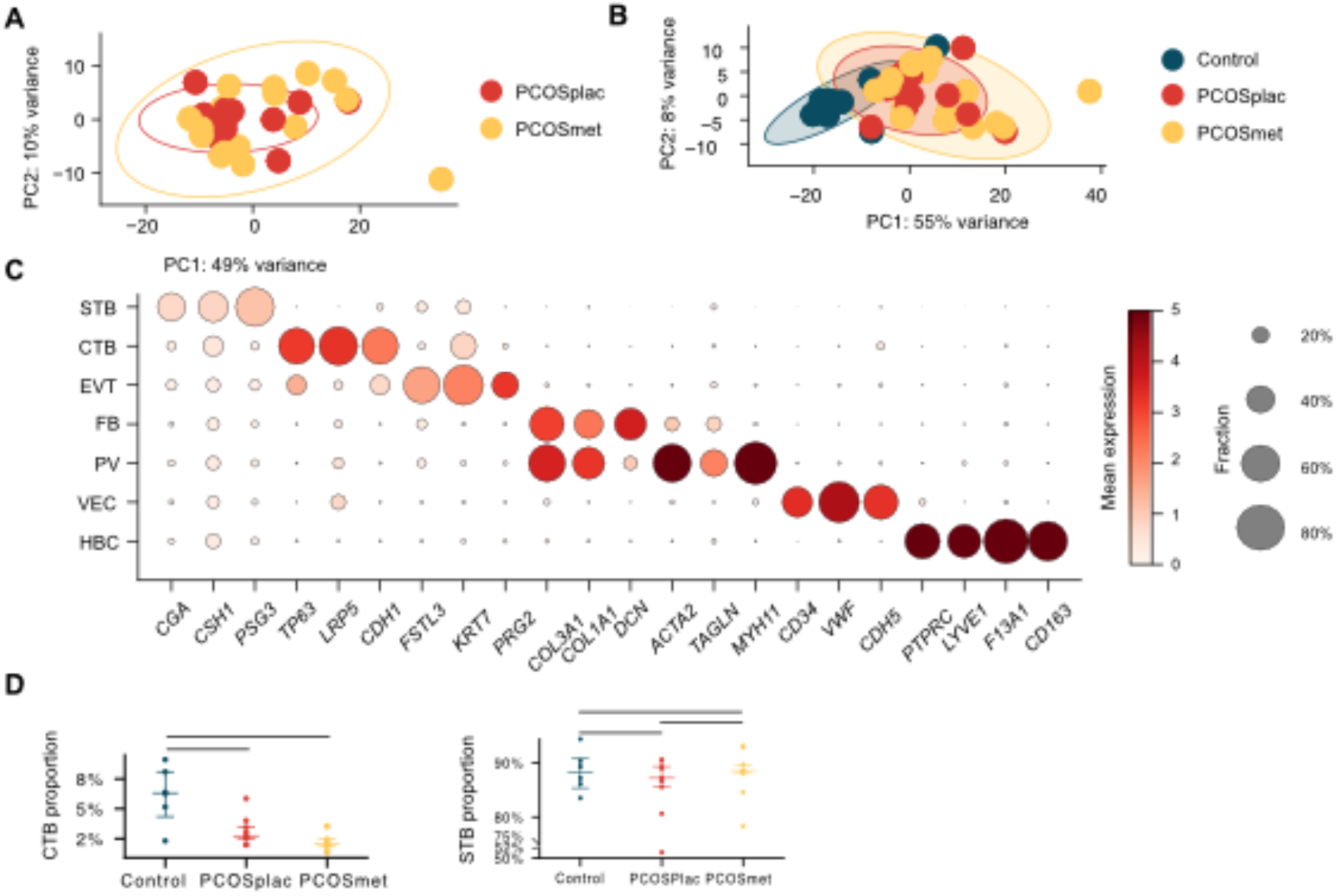
Transcriptional landscape, marker expression, cell composition, and differential gene patterns in PMOS placentas. **(A)** Principal component analysis (PCA) of the transcriptional profiles of placentas from women with PMOS (PMOSplac, PMOSmet) at the bulk level. **(B)** PCA of placenta samples from PMOSplac, PMOSmet, and Control. **(C)** Dot plots showing the expression patterns of marker genes. **(D)** scCODA^28^ test of CTB and STB proportion.

**Fig. S2.**
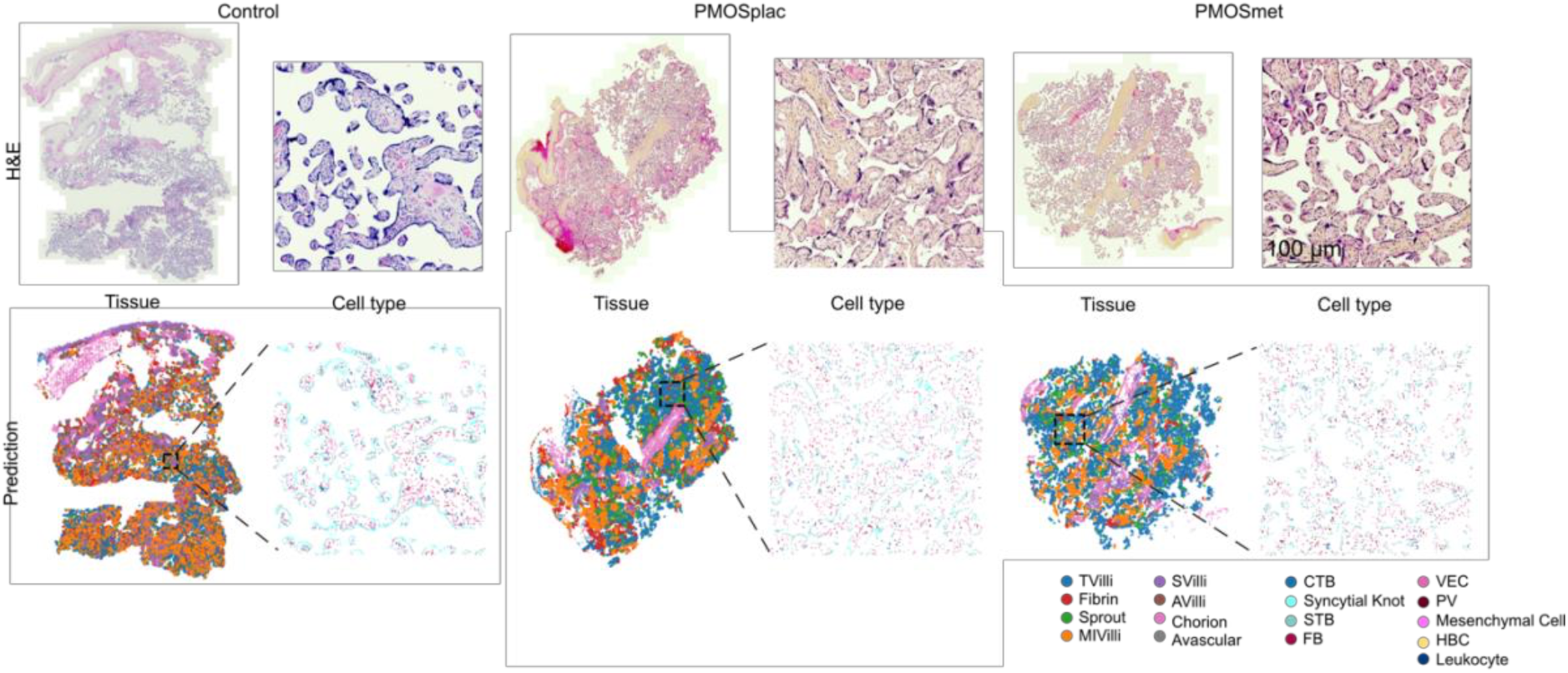
Tissue types and cell types of selected samples from Control and PMOS groups inferred by HAPPY. (Vanea et al., 2024). TVilli: Terminal villi, Sprout: Villous sprout, MIVilli: Mature intermediate villi, SVilli: Stem villi, AVilli: Anchoring villi, Chorion: Chorionic plate, Avascular: Avascular villi.

**Fig. S3.**
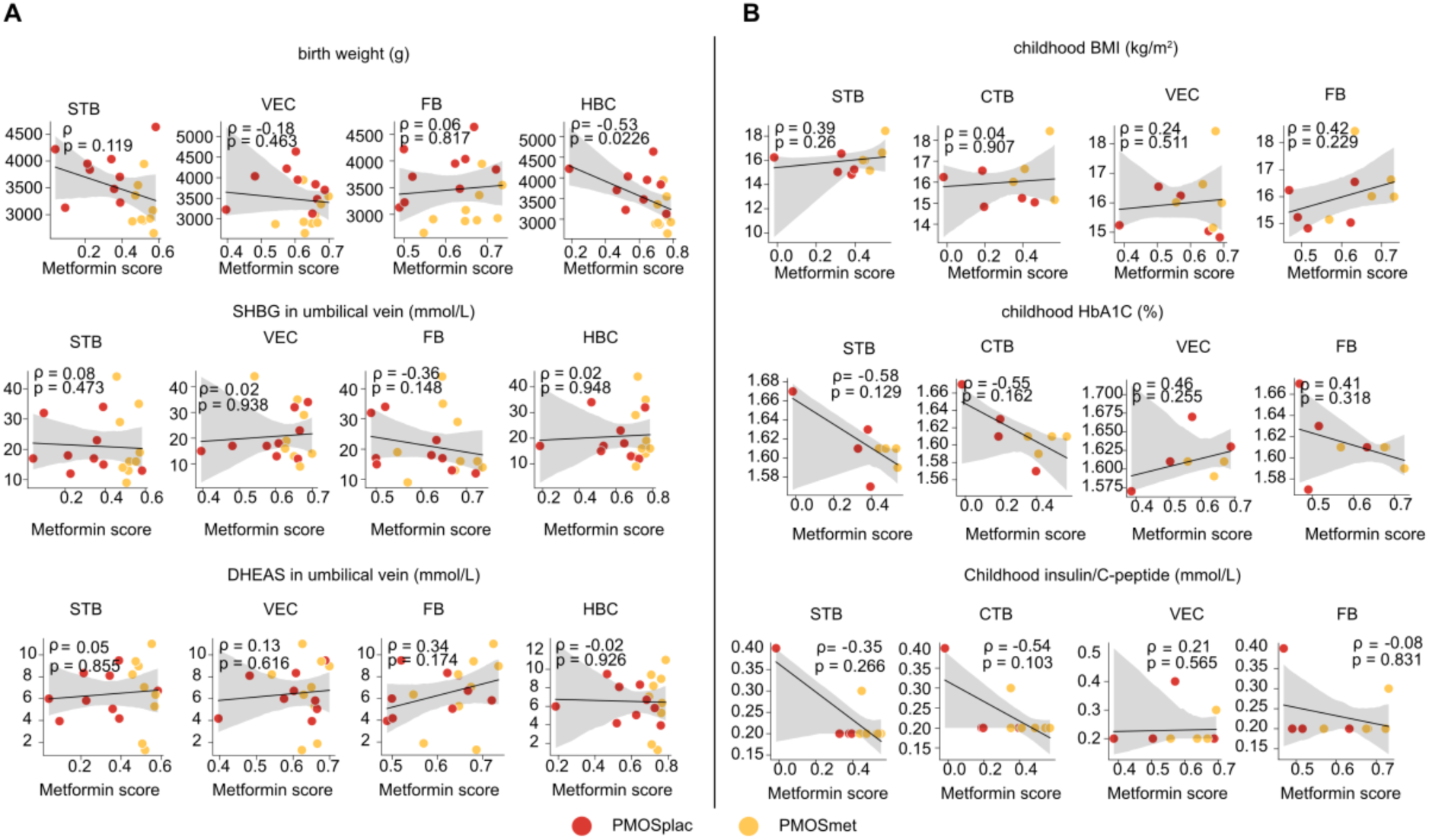
Neonatal and childhood biomedical measurements of offspring born to women with PMOS. **(A)** Correlation of metformin score and birth weight, SHBG and DHEAS levels in umbilical vein. And the metformin score, i.e., score of metformin-associated transcriptional alteration was done by Gene Set Variation Analysis (GSVA) using the DEGs between PMOSmet and PMOSplac in STBs, VECs, FBs, and HBCs as gene sets. The line is the linear regression fit and the shaded band is its 95% confidence interval. **(B)** Correlation of metformin score and childhood body mass index (BMI), insulin/C-peptide and HbA1C levels. And the metformin score, i.e., score of metformin-associated transcriptional alteration was done by GSVA using the DEGs between PMOSmet and PMOSplac in CTBs, STBs, VECs, and FBs as gene sets.

**Fig. S4.**
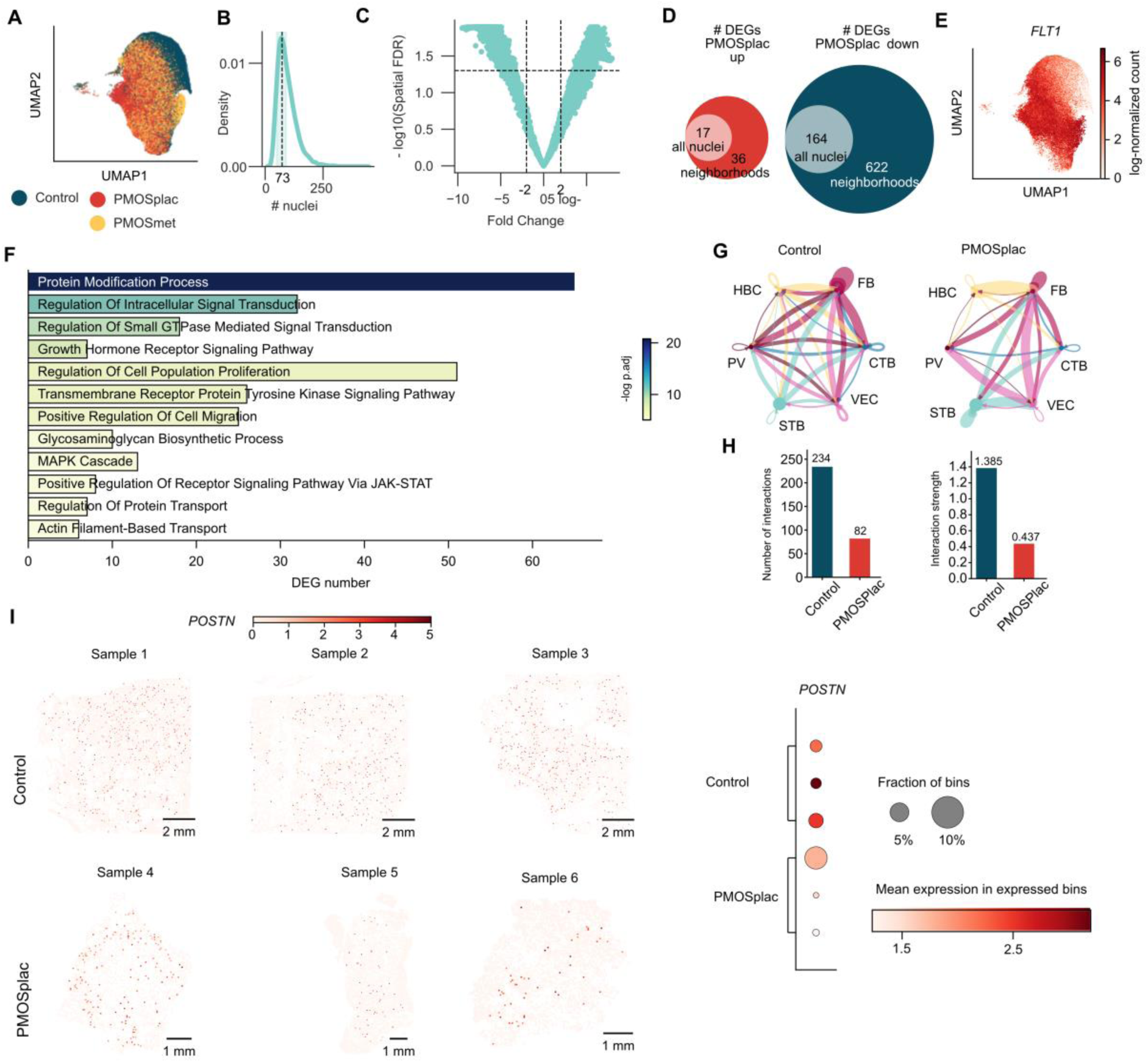
Differential expression analyses of STBs and CTBs, and general cell-cell communication inference across cell types in placental villi. **(A)** UMAP of STB nuclei colored by group. (B) Distribution of number of nuclei in neighborhoods. (C) Differential abundance of PMOSplac and Control in each neighborhood. (D) Venn-diagram of up- and down-regulated genes detected with all nuclei or nuclei in enriched neighborhoods between PMOSplac and Control. (E) UMAP of STB nuclei colored by *FLT1* expression. (F) Barplot showing the top 10 enriched gene ontology terms of the down-regulated genes in STBs of PMOSplac compared with Control. The terms are ranked by adjusted p-values of hypergeometric test. (G) Circle plot showing the strength of ligand-receptor interactions between pairwise cell populations among the major cell populations in Control and PMOSplac groups. The edge width was proportional to the strength of L-R pairs. **(H)** Comparison of cell–cell communication between Control and PMOSplac. (Left) Number of inferred interactions. (Right) Interaction strength. I. (Left) Spatial scatter plot of *POSTN* expression. (Right) Dotplot summarizes the fraction and mean expression of *POSTN* in expressed bins.

**Fig. S5.**
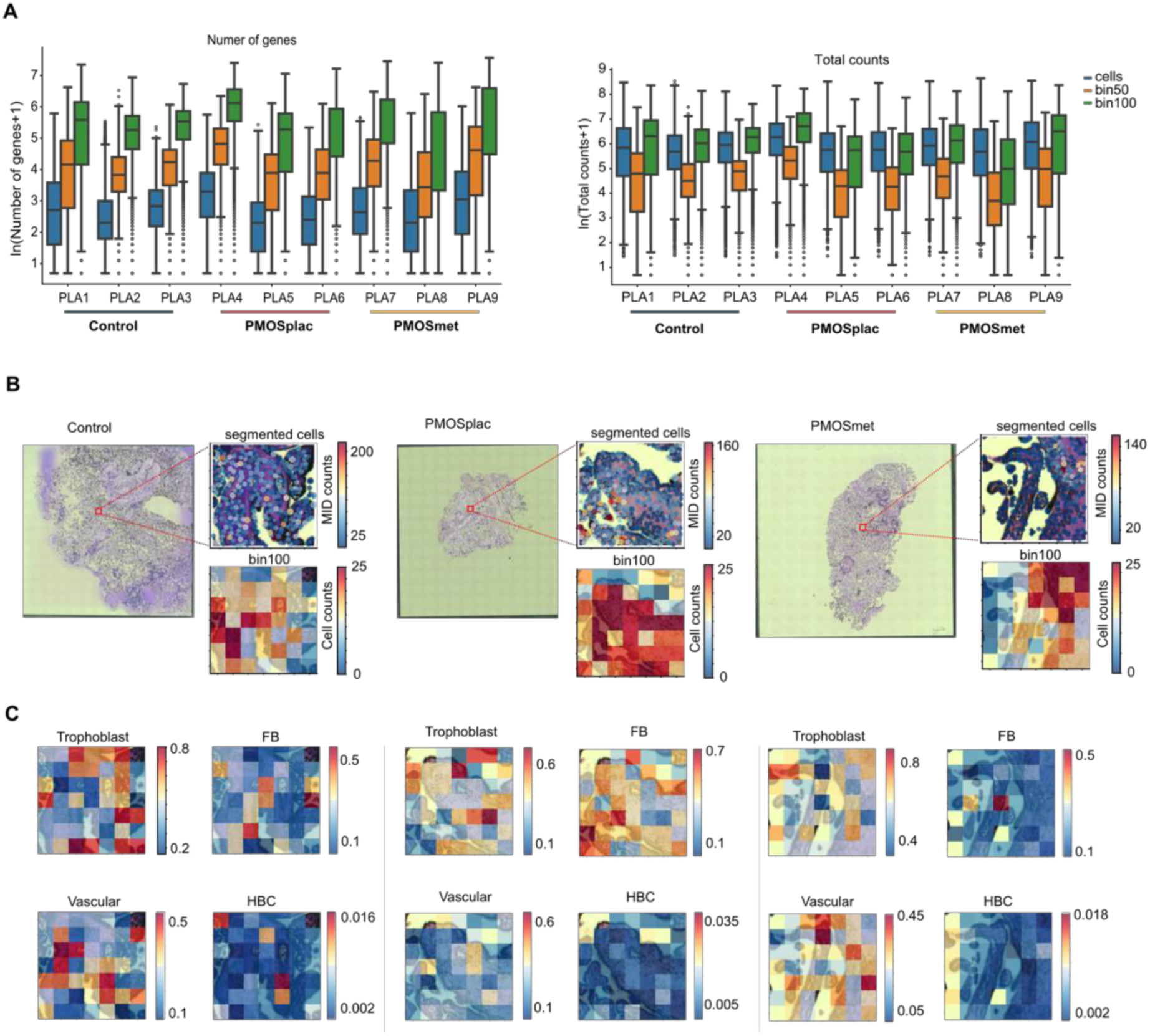
Quality control and spatial visualization of deconvoluted cell types of Stereo-seq data. **(A)** Boxplots showing number of genes and total counts detected at single-cell, bin50, and bin100 pseudo-spot level. n=3 placentas per group. **(B)** Spatial visualization single-cell with detected molecular identifiers (MID) counts and bin100 pseudo-spot with cell counts. **(C)** Cell type deconvolution on bin100 pseudo-spots. Proportion of mapping (color bar) of major cell types in cropped regions as in **B.**

**Fig. S6.**
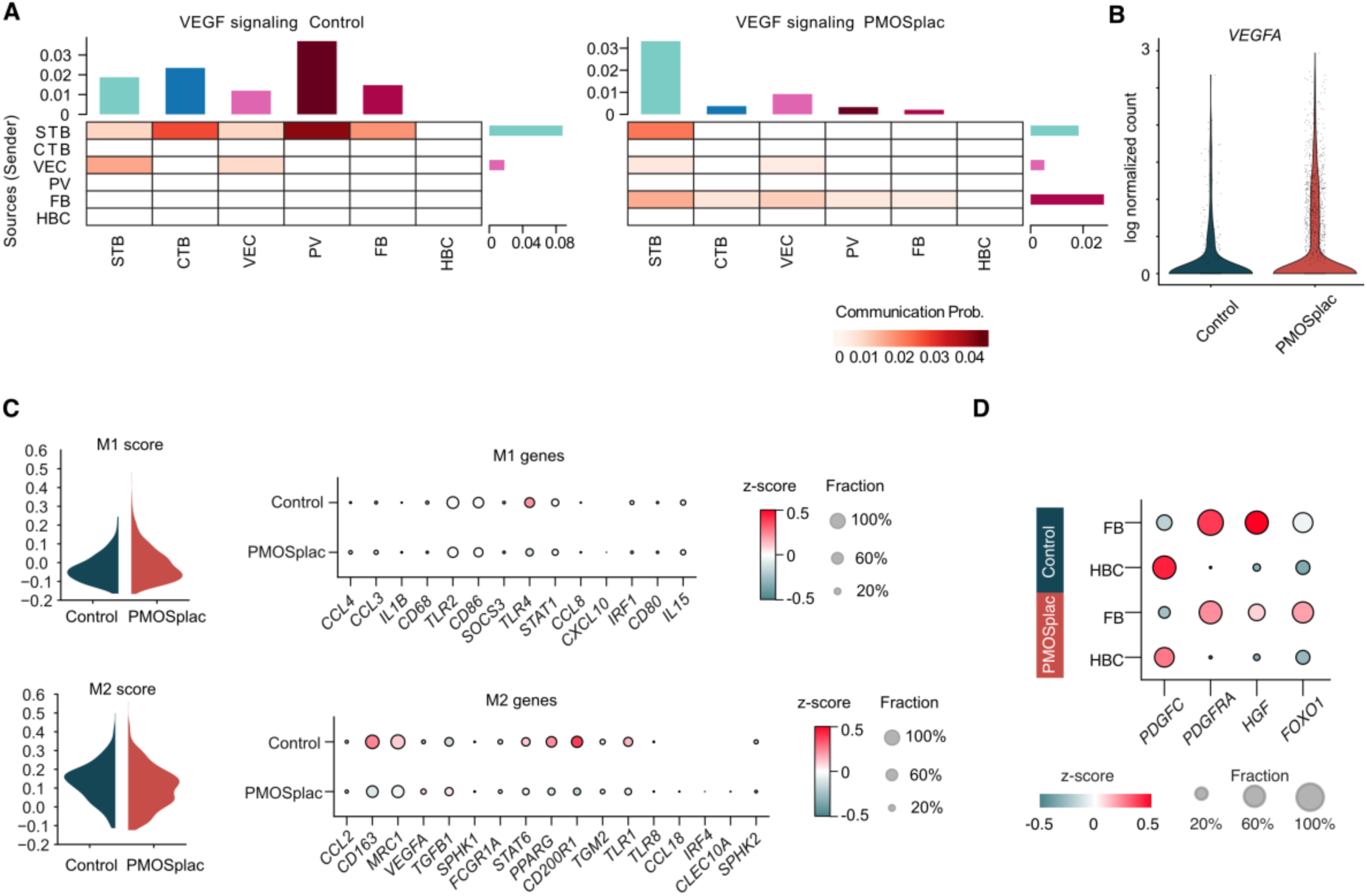
Stromal cells in PMOS placenta. **(A)** Gene set expression scores from the genes related to HBCs states in Control and PMOSplac. (Top) M1, (Bottom) M2. **(B)** Violin plot showing VEGFA expression in FBs. **(C)** Heatmap showing the VEGF signaling strength of the inferred cell-cell communication network in Control and PMOSplac samples. **(D)** PDGF signaling related gene expression in HBC and FB.

